# Inhibition of JAK-STAT pathway corrects salivary gland inflammation and interferon driven immune activation in Sjögren’s Disease

**DOI:** 10.1101/2023.08.16.23294130

**Authors:** Sarthak Gupta, Eiko Yamada, Hiroyuki Nakamura, Paola Perez, Thomas J.F. Pranzatelli, Kalie Dominick, Shyh-Ing Jang, Mehdi Abed, Daniel Martin, Peter Burbelo, Changyu Zheng, Ben French, Ilias Alevizos, Zohreh Khavandgar, Margaret Beach, Eileen Pelayo, Brian Walitt, Sarfaraz Hasni, Mariana J. Kaplan, Mayank Tandon, M. Teresa Magone, David E. Kleiner, John A. Chiorini, Alan N. Baer, Blake M. Warner

## Abstract

**Objectives:** Inflammatory cytokines that signal through the JAK-STAT pathway, especially interferons (IFNs), are implicated in Sjögren’s Disease (SjD). Although inhibition of JAKs is effective in other autoimmune diseases, a systematic investigation of IFN-JAK-STAT signaling and effect of JAK inhibitor (JAKi) therapy in SjD-affected human tissues has not been reported.

**Methods:** Human minor salivary glands (MSGs) and peripheral blood mononuclear cells (PBMCs) were investigated using bulk or single cell (sc) RNA sequencing (RNAseq), immunofluorescence microscopy (IF), and flow cytometry. *Ex vivo* culture assays on PBMCs and primary salivary gland epithelial cell (pSGEC) lines were performed to model changes in target tissues before and after JAKi.

**Results:** RNAseq and IF showed activated JAK-STAT pathway in SjD MSGs. Elevated IFN-stimulated gene (ISGs) expression associated with clinical variables (e.g., focus scores, anti-SSA positivity). scRNAseq of MSGs exhibited cell-type specific upregulation of JAK-STAT and ISGs; PBMCs showed similar trends, including markedly upregulated ISGs in monocytes. *Ex vivo* studies showed elevated basal pSTAT levels in SjD MSGs and PBMCs that were corrected with JAKi. SjD-derived pSGECs exhibited higher basal ISG expressions and exaggerated responses to IFNβ, which were normalized by JAKi without cytotoxicity.

**Conclusions:** SjD patients’ tissues exhibit increased expression of ISGs and activation of the JAK-STAT pathway in a cell type-dependent manner. JAKi normalizes this aberrant signaling at the tissue level and in PBMCs, suggesting a putative viable therapy for SjD, targeting both glandular and extraglandular symptoms. Predicated on these data, a Phase Ib/IIa randomized controlled trial to treat SjD with tofacitinib was initiated.

**What is already known on this topic?:**

- Upregulation of interferons (IFNs) has been reported in patients with SjD; however, a systematic investigation of their role at a cellular and tissue level in humans is lacking.

**What this study adds?:**

- Our findings conclusively show that the IFN-JAK-STAT pathway is activated in the salivary glands and PBMCs in patients with SjD
- Specific cells in the MSGs (infiltrating lymphocytes, epithelial, antigen presenting cells, and endothelial cells) and in PBMCs (monocytes, NK cells, and dendritic cells) drive this IFN signature.
- We pinpoint cells responsive to JAK inhibition and illustrate in patient tissues that JAK inhibitors may be beneficial in SjD by uncoupling the pathogenic cytokine milieu and resultant epithelial tissue damage and dysfunction central to SjD.

**How this study might affect research, practice, or policy?:**

- SjD lacks an approved, efficacious and targeted therapy. Several large clinical trials have been unsuccessful due in part to a lack of biologically relevant endpoints or predictive biomarkers. We establish a multimodal testing platform using human tissues from SjD patients to identify actionable targets and to directly test treatment effects. Our data suggest that blocking the IFN-JAK-STAT pathway by using JAKi is a rational therapy for SjD. Moreover, these data can also serve as biological endpoints for clinical trials [NCT04496960].

## Introduction

Despite being the second most common systemic autoimmune rheumatic disease, Sjögren’s Disease (SjD) lacks a precise etiology or an approved and efficacious therapy that meaningfully manages symptoms or alters disease progression.^1^ Concerted efforts have been expended toward identifying the pathogenic mechanisms of SjD; these efforts have led to the development and testing of many medications to treat SjD.^2^ However, few agents met their primary endpoints until very recently, and only limited biological data accompany these studies.^2^

One of the reasons for these limitations could be the heterogeneous clinical presentation of patients with SjD. Chronic inflammatory lymphocytic infiltration of the exocrine glands leading to dysfunction and ultimate destruction of the tissue is characteristic of SjD. Most SjD patients present with symptoms involving the salivary and lacrimal glands. Other tissues that can be affected include: the skin; the respiratory tract nervous system, kidneys, and the vagina, along with systemic symptoms including fatigue, widespread musculoskeletal pain, and polyarthritis.^34^

Many of the inflammatory cytokines implicated in SjD pathogenesis, in particular Type-I and Type-II interferons (IFNs), interleukins (IL)-6, IL-7, IL-12, and IL-21, signal through the Janus Kinases (JAK)-Signal Transducer and Activator of Transcription (STAT) pathway.^5^ However, a comprehensive understanding of IFN signaling and the involvement of the JAK-STAT pathway in SjD tissues is lacking. Clarification of cell types driving disease pathogenesis in the peripheral blood and in the salivary glands will facilitate the development of targeted therapies for SjD.

In this study, we aimed to characterize JAK-STAT pathway utilization in SjD-affected tissues and establish a human-tissue-based experimental rationale for examining the efficacy of JAK inhibitors (JAKi) in SjD participants. We used an ‘omics’ platform to identify high-potential tractable pathways in the affected tissue compartments in SjD. We first performed ‘bulk’ RNA sequencing (RNAseq) on minor salivary glands (MSGs) from patients with SjD and demonstrated activation of the JAK-STAT pathway. These findings were corroborated using immunofluorescent staining (IF) of salivary glands, and flow cytometry analysis of the salivary glands and peripheral blood mononuclear cells (PBMCs) from patients with SjD. Using single cell RNAseq (scRNAseq), we were able to identify cells in the glands and peripheral blood that upregulate IFNs through the JAK-STAT pathway in MSGs and then confirmed these findings using flow cytometry and *ex vivo* culture assays. Finally, we used patients’ tissues and primary cell models to show that JAKi can correct this altered pathway activation without cytotoxicity as a high-potential targeted therapy for SjD.

## Materials and Methods

### Subjects and ethical approval

Study subjects provided informed consent prior to the initiation of any study procedure, and they were evaluated and classified comprehensively according to 2016 American College of Rheumatology (ACR) and the European League Against Rheumatism (EULAR) classification criteria.^6^ Comparator group included subjects who did not meet 2016 ACR-EULAR criteria (non-SjD) or healthy volunteers (HV).^7^ All subjects were screened for evidence of systemic autoimmunity and received comprehensive oral/sialometric, rheumatological, and ophthalmological investigations. Clinical investigations were conducted in accordance with the Declaration of Helsinki principles. All studies using human samples were approved by the NIH IRB (15-D-0051, NCT00001390; 11-D-0172, NCT02327884, or 94-D-0018, NCT00001196; PI-Warner).

### Patient and Public Involvement

Patients or the public were not involved in the design, or conduct, or reporting, or dissemination plans of our research.

### Human Salivary Gland RNA Sequencing

Bulk RNA sequencing was performed as previously described.^8^ RNAseq data that passed quality control was deposited in dbGaP: phs001842.v1.p1.

### Single Cell RNA Sequencing of the MSGs and PBMCs

MSG biopsies and PBMCs were processed for scRNAseq as previously described (**supplemental table 1**).^9,10^ scRNAseq data sets were analyzed in Python using Scanpy.

### Immunofluorescence microscopy and analysis on MSGs

*Detection of JAK1 and JAK3 in MSG*. FFPE MSG sections were processed following standard procedures. Sections were blocked, incubated in antibodies, and mounted (**supplemental table 2**). *Immunofluorescence in primary cell culture assays.* Primary salivary gland epithelial cells (pSGECs) were plated on chamber slides and stimulated as indicated, then stained with antibodies (**supplemental table 2**). Images were acquired on Nikon A1 HD (Nikon) confocal microscope and processed with CellProfiler in ImageJ (Broad Institute).^11^

### Flow cytometry

Freshly biopsied MSGs were dissociated as described above. Multicolor flow cytometry was used to quantify the phosphorylation status of pSTATs in gated cell subset populations (**supplemental table 2** and **supplemental method 1, 2**).

### Assessment of serum proteome

Proteomic profiles were measured in serum (50μL) using the SOMAscan Assay V1.3 (SomaLogic, Inc.) at the Trans-NIH Center for Human Immunology, Autoimmunity, and Inflammation, National Institutes of Health as previously reported.^12^

### RNA isolation and RT-qPCR

Standard Taqman assays (**supplemental table 3**) were performed to measure relative gene expression.

### Statistical Analysis for Difference in Immune Cell Populations/Cytokines

Statistical methods were employed using GraphPad Prism (GraphPad), MATLAB, or –R as described, and the type and nature of the data were considered when assessing differences in mean values and variances across biological and experimental replicates. A *p*-value of <0.05 was considered statistically significant.

## Results

### Genes in the JAK-STAT and Type-I IFN pathways are upregulated in MSGs from patients with SjD

To understand the transcriptome wide impacts of SjD on the MSGs, we first performed ‘bulk’ RNAseq on SjD and HV MSGs (**figure 1A**). Unsupervised clustering generally segregated SjD from HV MSGs based on differentially expressed genes (DEGs) (**figure 1B, C**). MSGs from SjD subjects were transcriptionally more active. Immune pathway genes, including ISGs (e.g., *IFI44L, IFI44, MX1, CXCL13*), were upregulated in SjD whereas canonical salivary genes were downregulated (**figure 1B, C**). Pathway enrichment analysis identified the *JAK-STAT Pathway* as one of the top significantly and differentially utilized pathway in SjD (**figure 1D**). Gene enrichment analysis showed consistent and direct regulation of the JAK-STAT pathway signaling through *IL7/IL15/IL21* via *JAK3* (**supplemental figure 1B**). We examined the mRNA expression of JAK genes and found that *JAKI* had the highest expression in all MSGs whether they were from SjD or HV. However, when comparing MSGs from SjD to those from HV, *JAK2* and *JAK3*, but not *JAK1* or *TYK2,* expression was differentially increased (**supplemental figure 1C**).

**Figure 1:**
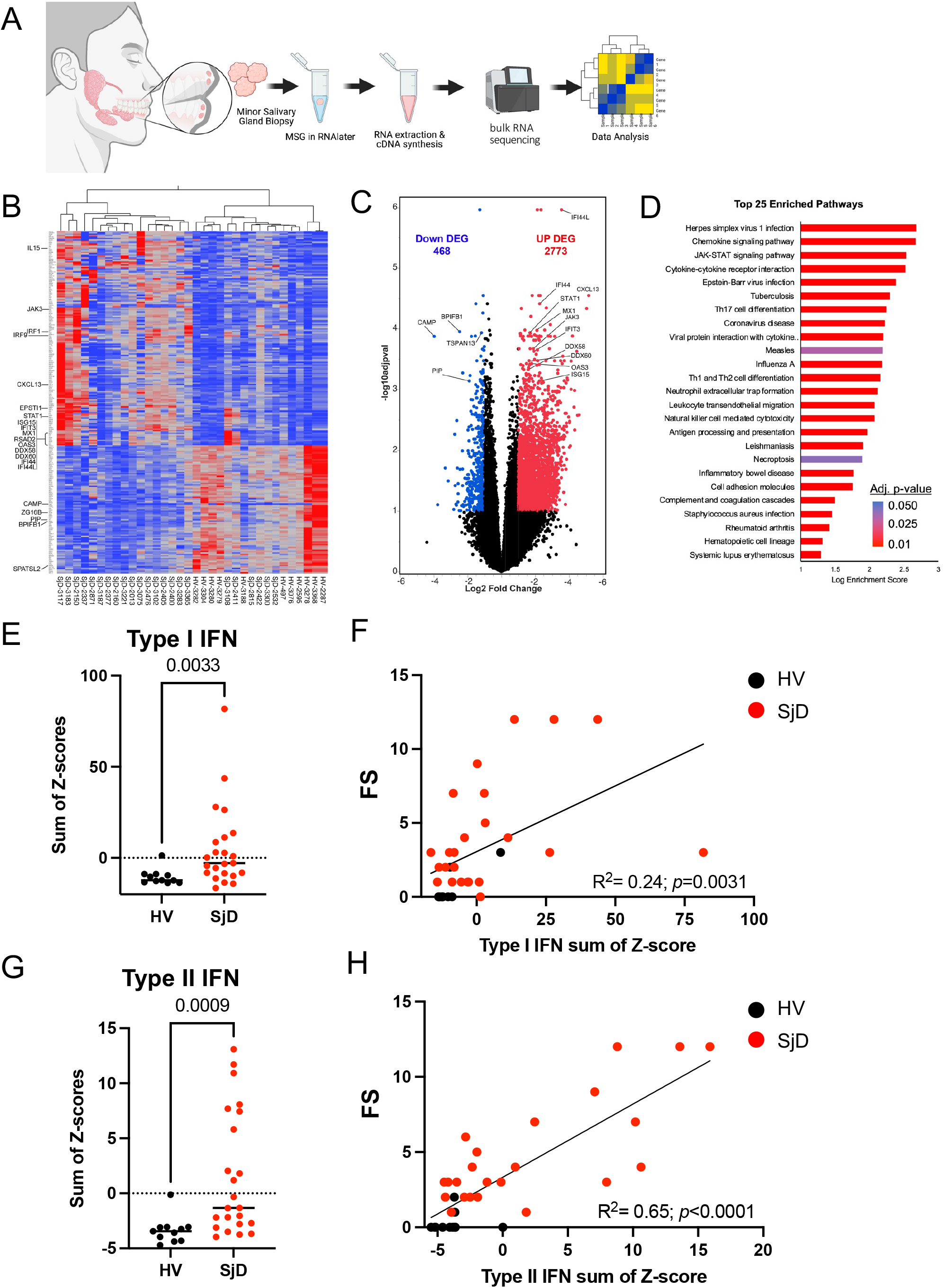
Bulk sequencing of minor salivary gland and IFN signature. (A) Overview of MSG biopsy and whole transcriptomic analysis using RNAseq from 22 SjD and 11 HV. (B) Heatmap illustrating the top 250 DEGs in MSG between SjD and HV highlighting multiple ISGs, cytokines, and interleukins, in the DEG. Immune pathway genes, including ISGs (e.g., *IFI44L, IFI44, MX1, CXCL13*), were upregulated whereas canonical salivary genes (e.g., *BPIFB2, PIP, ZG16B*) were down regulated in SjD. (C) Similarly, a volcano plot showing the DEGs between SjD and HV, in which some representative genes were highlighted. MSGs from SjD subjects were transcriptionally more active with 2773 upregulated versus 468 downregulated genes. (D) Pathway enrichment analysis identified JAK-STAT Pathway as one of the top three differentially utilized pathway amongst the 25 significantly enriched pathways in SjD at a *p*-adj<0.01 (E, G) Calculated Type-I and Type-II IFN scores revealed 2 through 2.5-fold mean increases in SjD MSGs compared to HV. Differences in mean values were compared using the Mann-Whitney U-test at a *p* < 0.05 deemed significant. (F, H) The activated IFN signature noted in our bulk RNAseq positively correlated with FS in the glands. Spearman correlation analysis was used to assess the significance between correlated values at a *p* <0.05. MSG, minor salivary gland; RNAseq, RNA sequencing; SjD, Sjogren’s Disease; HV, healthy volunteer; DEGs, differentially expressed genes; ISG, interferon stimulated genes; FS, focus score.

Although the bulk RNAseq was unable to reliably detect Type-I IFN transcripts, these data demonstrated increased *IFNG* (**supplemental figure 1D**). Thus, we employed surrogate readouts of Type-I and Type-II IFN gene expression using validated composite Type-I (21-gene)^13,14^ and Type-II (8-gene)^15–17^ IFN scores to demark patients with enhanced JAK-STAT pathway signaling. Calculated Type-I and Type-II IFN scores^18^ revealed 2-fold and 2.5-fold mean increases, respectively, in SjD patients’ MSGs compared to HV glands (*p=0.0033* and *p=0.0009*, respectively; **figure 1E, G**). Of the 24 SjD subjects, 17 (71%) had elevated Type-I or Type-II ISG scores; 7 exhibited elevations in both Type-I and Type-II IFN ISGs; while 5 each exhibited elevated Type-I or Type-II ISGs. Seven did not show significant elevation of either and were enriched for anti-SSA antibody negative subjects (5/7) (**figure 1E, G**). Type-I and Type-II ISG scores exhibited modest, positive, statistically significant correlation, but not all the variance was explained.

Increased lymphocytic infiltration, quantified as a focus score (FS), is an independent predictor of deterioration of exocrine gland function.^19^ SjD is related to both loss of epithelial cells and greater inflammation in the glands. The Type-I and Type-II IFN scores from our bulk RNAseq positively correlated with FS in the glands; however, Type-II IFN exhibited a stronger correlation than Type-I IFN (*R^2=0.24, p=0.0031* and *R^2=0.65, p<0.0001*, respectively; **figure 1F, H**). Our data suggests that Type-II IFN signature serves as a surrogate for IFNγ produced by T cells in the glands; while Type-I IFN signaling is appreciably more disease-specific and integrates the contributions of the epithelial cells, dendritic cells, and monocytes in the MSG. Moreover, it is well-documented that there is positive crosstalk between Type-I IFNs (α and β) and Type-II IFN (γ) receptor signaling. In our cohort most patients exhibited elevated Type-I signaling. Thus, we focused on Type-I signaling in subsequent experiments, acknowledging that signaling *in vivo/in situ* is far more complex.

These findings confirm that SjD pathogenesis involves IFN-JAK-STAT pathway. However, “bulk” RNAseq does not precisely and robustly predict the compositional cell types or infer the cell states changes contributing to these phenomena in SjD-affected MSG.

### Expression of JAK1 and JAK3 in the salivary glands is cell type-specific

To pinpoint the cell types and expression levels of JAK1 and JAK3 in the MSG, we used IF, whole slide imaging and analysis, and tSNE visualization (**figure 2A**). These results revealed the expected cellular proportion changes in the MSGs. Keratin-18 expression was restricted to epithelial structures (+6%, *p<0.001*; **figure 2B, C** and **supplemental figure 2A-C**). Per-cell JAK1 expression was mildly elevated in immune cells in SjD, while per-cell JAK3 expression showed both SjD-specific increased per-cell expression in epithelial cells and infiltrating immune cells in SjD (+35%, *p*<0.0001 and +15%, *p*<0.0001, respectively; **figure 2C**). It is generally assumed that high IFN signaling is driven by infiltration of immune cells; however, the IF and RNAseq results independently confirm that signaling is directed in part by the involved epithelium.

**Figure 2:**
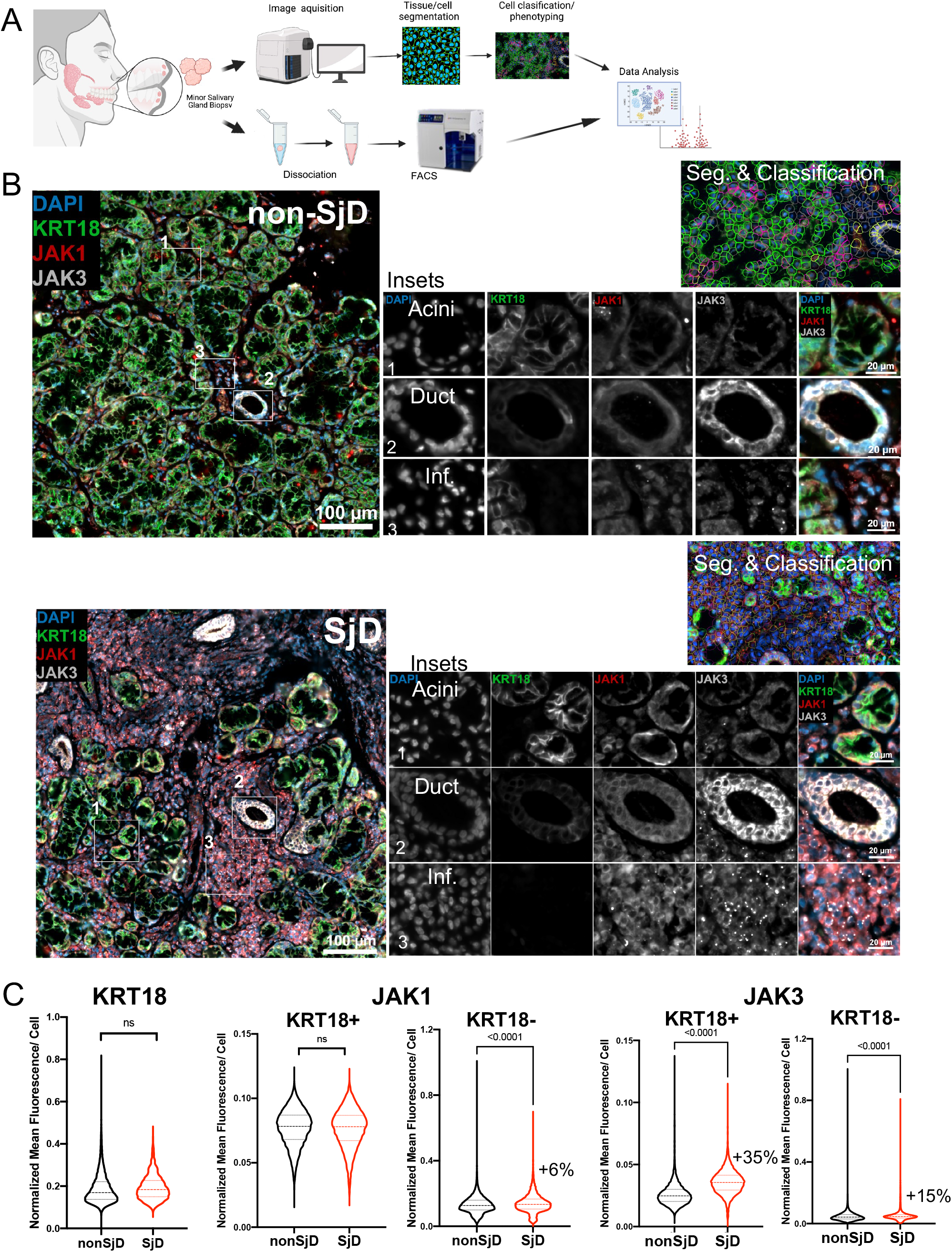
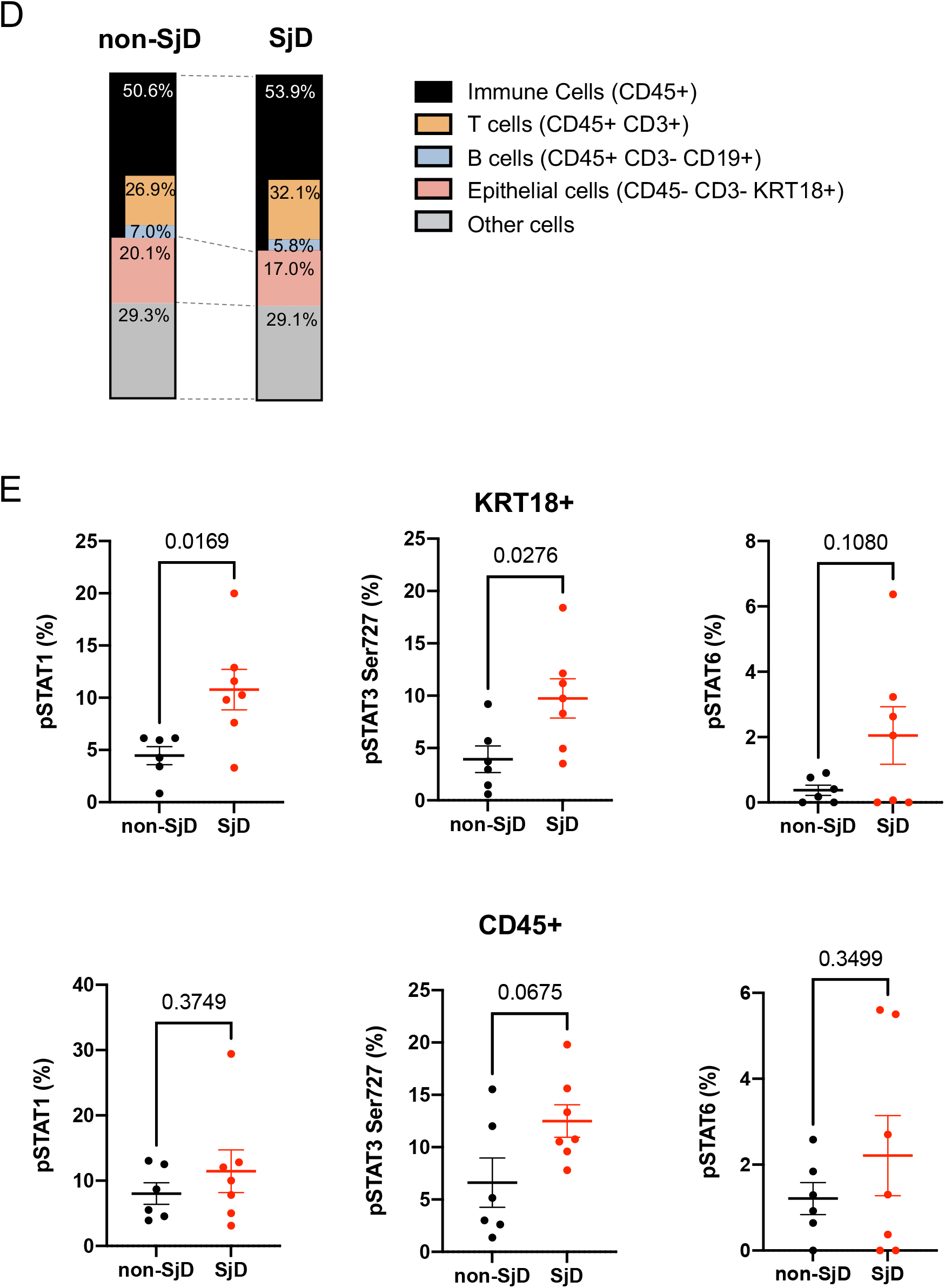
Immunofluorescence microscopy. (A) Overview of MSG biopsy, image acquisition and flow cytometry. (B) MSG IF image showing upregulated expression of JAK1 and JAK3 in SjD epithelial and infiltrating immune cells. JAK3 was especially enrich in ductal cells. Cellular proportion changes in the SjD MSGs showed greater numbers of immune cells, less numbers of epithelial cells. (C) Mean fluorescence of KRT18, JAK1 and JAK3 in SjD and non-SjD showing cellular proportion in the MSGs. Expression of JAK1 localized to immune cells (+6%; *p*<0.0001, Mann-Whitney Test) but JAK3 expression was seen in both epithelial and infiltrating immune cells (35% and 15%, respectively; *p*<0.0001, Mann-Whitney Test). (D) Cellular population change in 7 SjD and 6 non-SjD was characterized by flow cytometry showed reduced numbers of epithelial cells and increased numbers of infiltrating immune cells in SjD MSGs compared to controls. (E) Flow cytometry of MSG represented the frequency of pSTAT proteins: pSTAT1 (2.1-fold, *p*=0.017), pSTAT3(Ser727) (2.2-fold, *p*=0.031), and pSTAT6 (3.3-fold, *p*=0.112) were higher at baseline in SjD epithelial cells compared to non-SjD. Although not reaching the threshold of statistical significance, the frequency of pSTAT proteins (i.e., pSTAT3(Ser727) (*p*=0.056)) on CD45+ cells showed a similar frequency of pSTAT proteins in the SjD MSG. P value was calculated using Welch’s test. MSG, minor salivary gland; IF, immune fluorescent; SjD, Sjogren’s Disease; KRT18, Keratin-18.

### Increased phosphorylated STAT proteins in SjD MSGs confirm activation of JAK-STAT pathway

To directly confirm the activation status of the JAK-STAT pathway in MSGs, we measured the frequency of pSTAT proteins by flow cytometry in freshly biopsied and dissociated MSGs (**figure 2A**). In general, flow cytometry exhibited cellular proportion changes consistent with the IF results in the glands (**figure 2D**). The frequency of pSTAT1, pSTAT3(Ser727) and pSTAT6 proteins were higher at baseline in SjD epithelial cells compared to non-SjD (2.0-fold, *p=0.017*; 2.9-fold, *p=0.028*; and 6.9-fold, *p=0.1080*, respectively; **figure 2E**), directly supporting elevated activation of the JAK-STAT pathway in SjD epithelial cells. Moreover, the frequency of pSTAT proteins on immune cells showed a similar pattern in SjD-affected MSGs (1.4-fold, *p=0.3749*; 2.6-fold*, p=0.068*; and 1.2-fold, *p=0.3499*, respectively; **figure 2E**), with appreciably less significant differences.

### High IFN signature and activated JAK-STAT pathway in SjD MSGs is due to salivary gland epithelial and infiltrating immune cells

Our results implicate coordinated interactions between the epithelium and the immune infiltrate (i.e., autoimmune epithelitis).^3^ To better understand the transcriptional impact of SjD on each cell type, we then analyzed single cell transcriptomes from MSGs from SjD and non-SjD subjects. Leiden clustering of scRNAseq data identified 11 clusters of cells representing the general cell types in the MSGs (**figure 3A** and **supplemental figure 3A, C**). Expectedly, compared to non-SjD MSGs—and consistent with both the IF and flow cytometry results—the proportion of seromucous acinar cells was reduced and immune cell infiltration was increased in SjD (**figure 3B**). Examining the top ten DEGs in each of the cell types demonstrated that SjD cells were dominated by ISGs (**figure 3C**). A Type-I IFN score was calculated for all cells in SjD and non-SjD and was increased in all cells in SjD patients’ glands (**figure 3D**). Moreover, seromucous acinar cells showed increased expression of multiple *JAK* genes, while ductal cells had increased expression of *JAK3* and *TYK2* (**figure 3E** and **supplemental figure 3C, D**). These results independently confirm our bulk and proteomics data and suggest that disease-specific utilization of this pathway in unique cell types.

**Figure 3:**
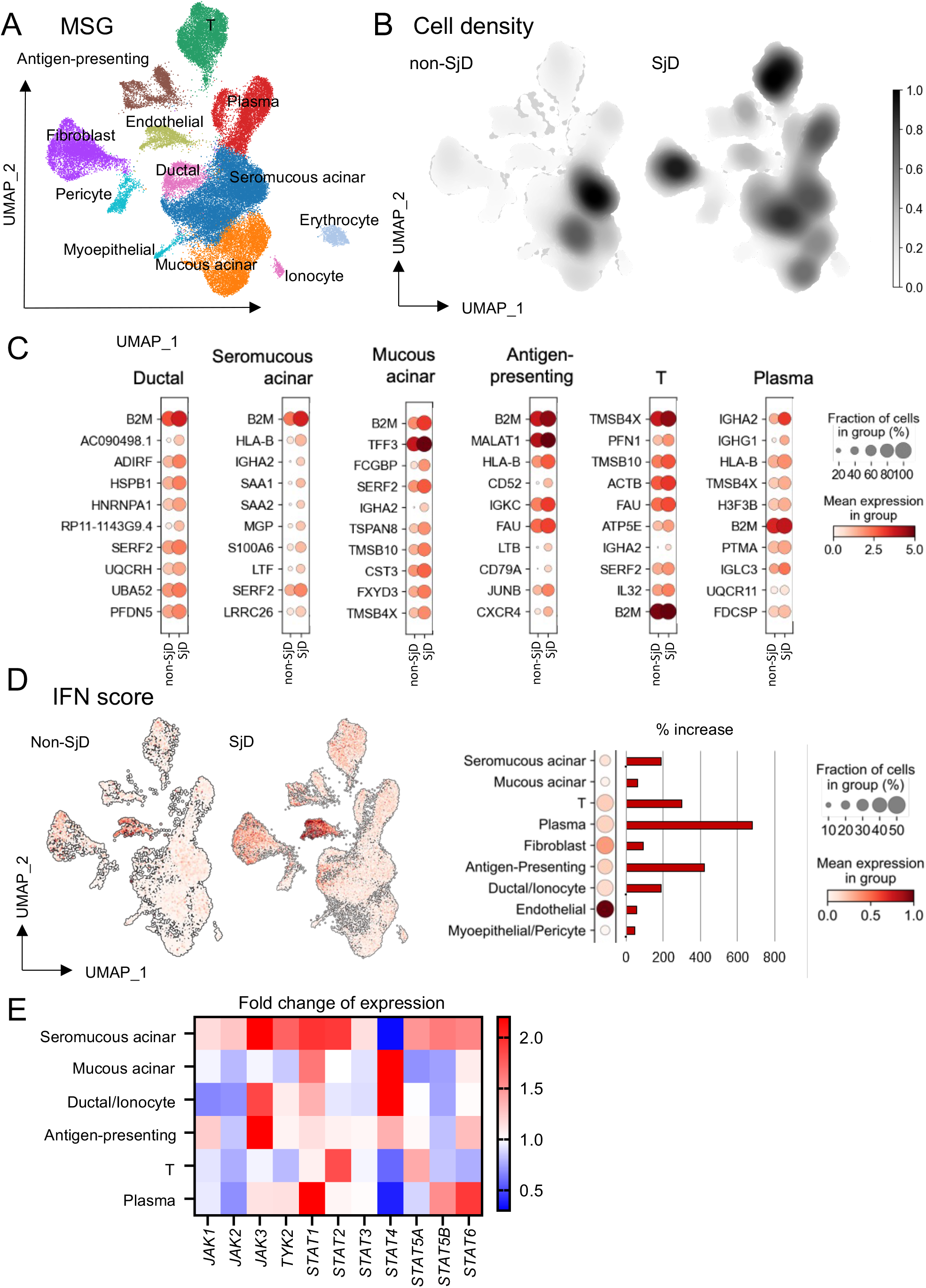
Single cell RNAseq and pSTATs frequencies of MSG. (A) UMAP embedding of the entire dataset colored by generated clusters labelled by cell type annotation. From all profiled MSG samples from 7 SjD and 5 non-SjD, Leiden clustering identified 11 different cell clusters corresponding to mucous (*MUC5B*) and seromucous acinar cells (*MUC7*), ductal cells (*S100A2*), plasma cells (*IGHA, IGHG1*), fibroblasts (*COL1A2*), myoepithelial (*KRT14*), pericytes (*ACTA2*), B-cells (*CD79A*), antigen presenting cells (*HLA-DRA, CD68*), T-lymphocytes (*CD3D*), and erythrocytes (*HBB*) (n=51736 cells). (B) Differential of cell density showing increased immune cell infiltration in SjD. (C) The top ten DEG in each of the cell types were dominated by ISGs including increased expression *B2M, HLA-B, SAA1, IL32,* and *MGP*. (D) Differential expression of IFN score in SjD and non-SjD, immune cells showed the biggest fold-changes in IFN score were in infiltrating immune cells (e.g., plasma cells: 700-fold, APCs: 400-fold; T cells: 300-fold; seromucous cells: 250-fold; ductal epithelial cells: 250-fold) exhibited higher IFN scores. (E) Fold change expression of JAK-STAT genes on all cell types. *JAK1* was the most ubiquitously expressed in MSGs and Seromucous acinar cells showed increased expression of all *JAKs.* UMAP, Uniform manifold approximation and projection; MSG, minor salivary gland; SjD, Sjogren’s Disease; DEG, differentially expressed genes.

### Sera from SjD have increased IFN Signature

As a prototypic systemic autoimmune disease, additional non-exocrine organs are also commonly affected in SjD. To estimate proteomic changes in the circulation, Somalogic aptamer-based 1.3K target proteomics was performed. Most proteins were significantly upregulated in sera from SjD patients compared to controls (**figure 4B**). Consistent with MSGs data, inflammation and IFN regulated proteins were upregulated in sera. A validated 4-protein IFN score demonstrated a significant increase of IFN-stimulated proteins in SjD patients’ sera compared to both non-SjD and HV sera (*p*<0.0001; **figure 4C**). These data indicate that IFN signatures in the glands are recapitulated in the blood and suggest similar pathogenic mechanisms affecting both tissue compartments.^2^

**Figure 4:**
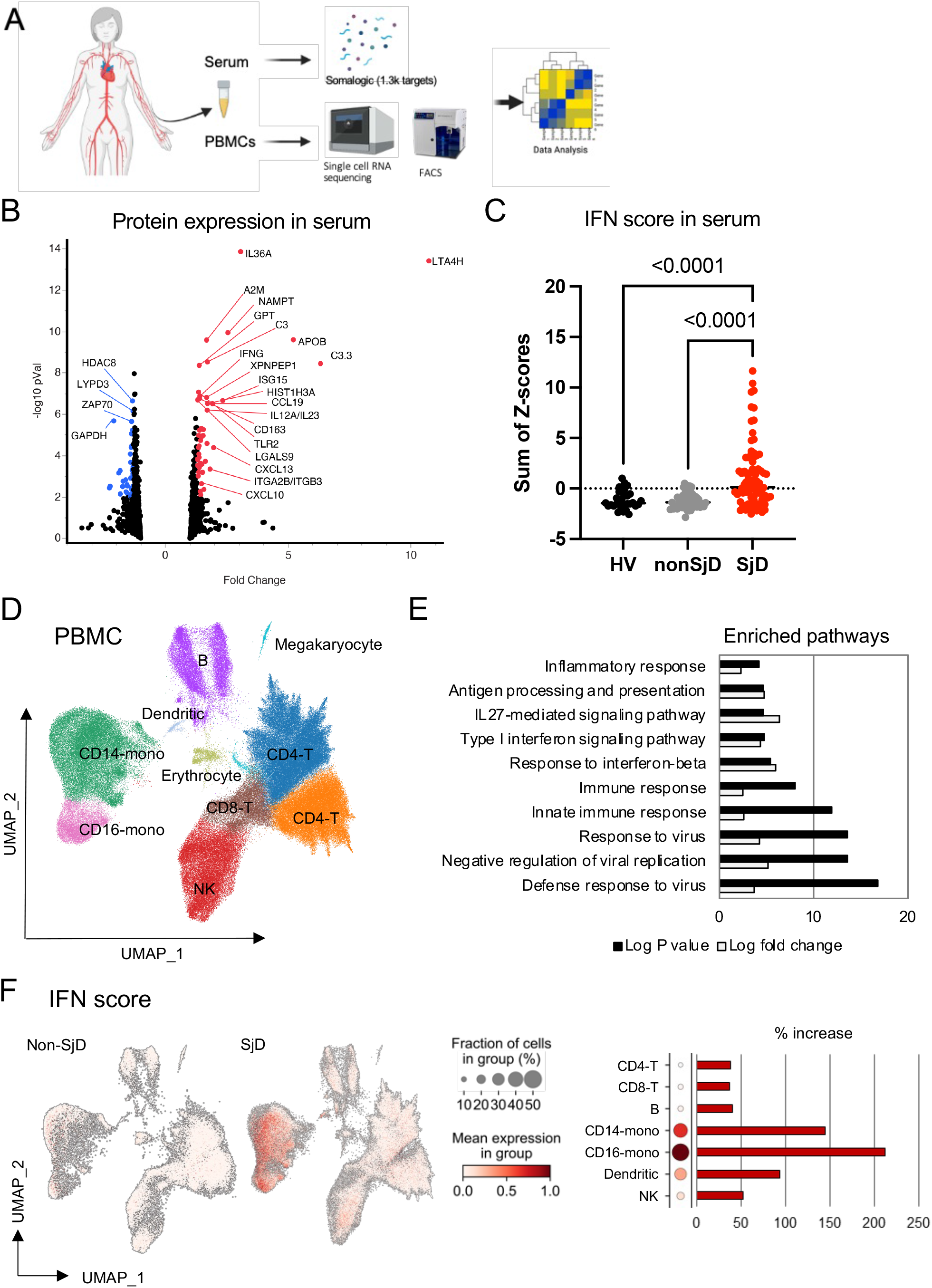
Single cell RNAseq and pSTATs frequencies of PBMC. (A) Overview of using patient’s serum and PBMC for several assays. (B) Somalogic aptamer-based 1.3K target proteomics analysis revealed most of proteins were significantly upregulated in serum from SjD in volcano plot showing protein expression, in which some representative genes were highlighted. (C) IFN regulated proteins were found to be upregulated in SjD serum (*p*<0.0001). P value was calculated using Kruskal-Wallis test. (D) UMAP embedding of the entire dataset colored by generated clusters labelled by cell type annotation. Leiden clustering identified 10 different cell clusters from all profiled PBMC samples from 8 SjD and 6 non-SjD (n=206687 cells). (E) The upregulated genes were associated with immune and inflammatory responses revealed by functional annotation analysis from PBMC scRNAseq. (F) Differential expression of IFN score, monocytes had the highest expression of ISGs, followed by dendritic cells and then NK cells. SjD, Sjogren’s Disease; UMAP, Uniform manifold approximation and projection; RNAseq, RNA sequencing; ISG, interferon stimulated genes.

### scRNA of PBMCs identify cells with activated JAK-STAT pathway

To better understand the transcriptional impact of SjD on each cell type in the peripheral blood, single-cell transcriptomes from PBMCs from SjD and non-SjD subjects were analyzed. scRNAseq identified 10 unique general types by Leiden clustering (**figure 4D** and **supplemental figure 4A, B**). DEG analysis showed upregulation of many ISGs (e.g., *IFI44L, IFIT3, ISG15, MX1,* and *IFI6*) in SjD PBMCs (**supplemental figure 4C**). Functional annotation analysis revealed that upregulated genes were mostly associated with immune and inflammatory responses, especially ‘*innate immunity against virus*’ and ‘*Type-I IFN responses*’ (**figure 4E**). Type-I IFN scores were projected on to each cell and were higher in SjD PBMCs compared to controls (**figure 4F**). Monocytes had the highest expression of ISGs in SjD, followed by dendritic cells, and then NK cells (**figure 4F**).

### Increased phosphorylated STAT proteins in SjD PBMCs confirmed activation of JAK-STAT pathway in blood

To directly estimate the activation of the JAK-STAT pathway in the blood, the frequencies of pSTAT proteins were measured by flow cytometry of PBMCs from patients with SjD and HVs. The frequencies of basal pSTAT proteins, pSTAT1, pSTAT3(Ser727) and pSTAT6, were higher in SjD patients compared to HVs (2.0-fold, *p=0.007*, 2.3-fold, *p=0.046*, and 1.3-fold, *p=0.044*, respectively; **figure 5** and **supplemental figure 5**). Of note, pSTAT3(Ser727), but not pSTAT3(Tyr705) was significantly upregulated in SjD patients. In aggregate, these findings indicate that enhanced Type-I IFN response in PBMCs could be involved in both the systemic aspect of SjD and local inflammation after infiltration into the MSGs. Our results suggest that we could employ this tissue as a surrogate model for assessing if identified drugs are biologically effective for SjD or stratify patients amenable to pathway targeting, *a priori*.

**Figure 5:**
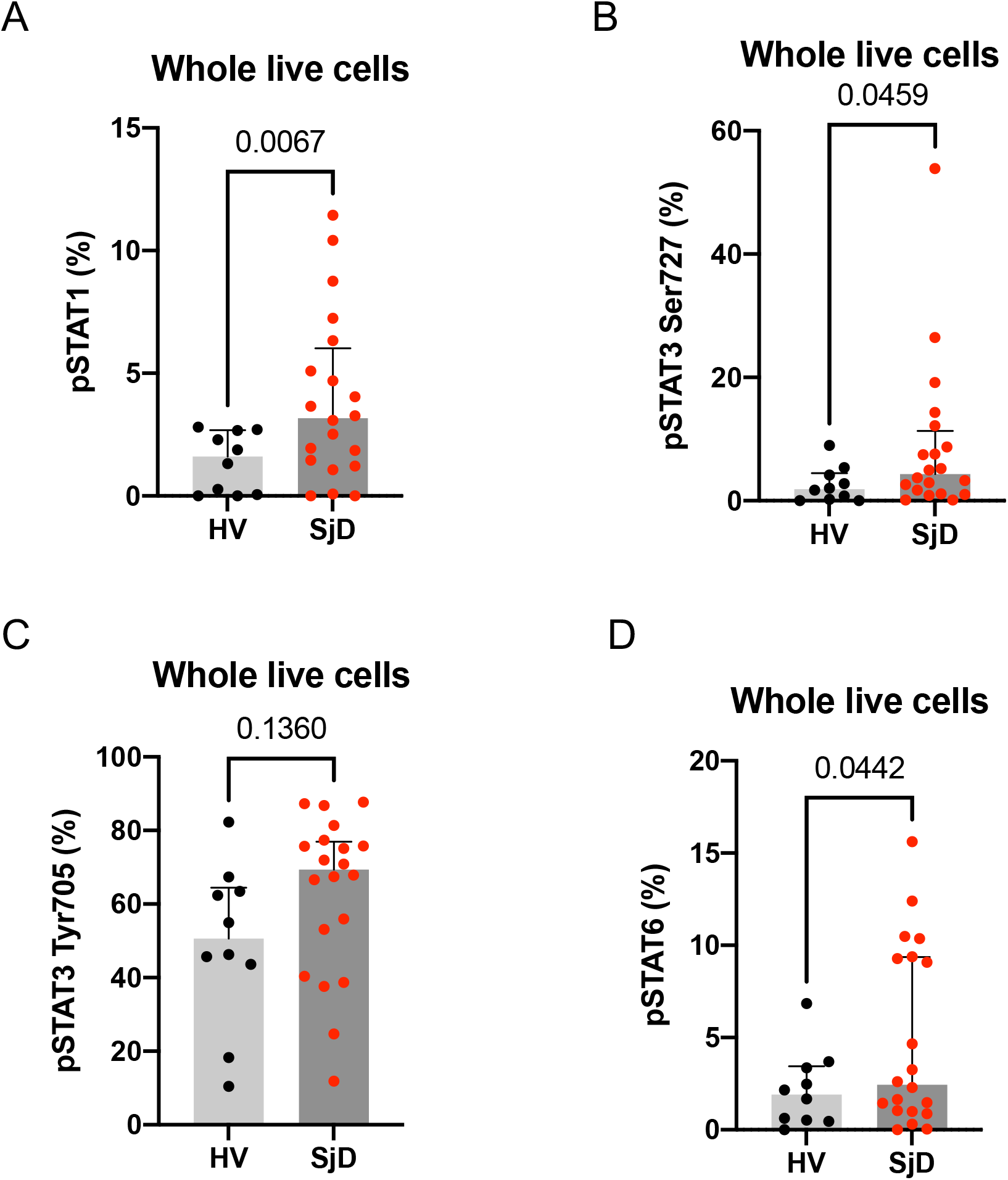
Basal pSTATs frequencies in PBMCs. (A-D) Flow cytometry analysis revealed basal pSTATs levels in PBMCs were upregulated in 21 SjD compared to 10 HV. P value was calculated using Welch’s test. SjD, Sjogren’s Disease; HV, healthy volunteer.

### Tofacitinib inhibits Type-I IFN response in PBMCs

Based on our transcriptomic and proteomic data, SjD pathogenesis may be selectively dependent on JAK3 and JAK1 in both immune and epithelial compartments. Thus, we tested the effectiveness of the JAK3/JAK1 semi-selective inhibitor, tofacitinib, in *ex vivo/in vitro* assays (**figure 6A**). Treatment with 5 μM tofacitinib blocked STATs phosphorylation status induced by IFNβ stimulation in PBMCs in all cell subsets without inducing cytotoxicity (**figure 6B** and **supplemental figure 6A, D**). scRNAseq showed that 5 μM tofacitinib downregulated 109 DEG, nearly all of which were ISGs, and no genes were upregulated. Tofacitinib suppressed the IFNβ-induced IFN signature to baseline levels (**figure 6C, D** and **supplemental figure 6B**). Cell clusters with sufficient number of representation of samples and cells in each treatment group cluster were analyzed for top DEG by pseudobulk analysis. T cells and monocytes exhibited greater numbers of downregulated ISGs in the context of tofacitinib inhibition. ISGs were among the top DEG in all subsets after IFNβ stimulation and tofacitinib normalized this response; cell type-specific effects were less dominant (**figure 6E**). A similar direction of effect was shown in PBMCs treated with 5μM tofacitinib without IFNβ stimulation, though there was appreciable inter-individual variability in responsiveness to tofacitinib (**supplemental figure 6C**). These data provide clear and direct evidence, using affected human tissues from SjD patients, that *in vitro* tofacitinib can abrogate basally activated JAK-STAT signaling and suggest that responsiveness to JAK inhibition is somewhat SjD-selective. These results raised the question as to whether a similar correction could be achieved in glandular epithelial cells.

**Figure 6:**
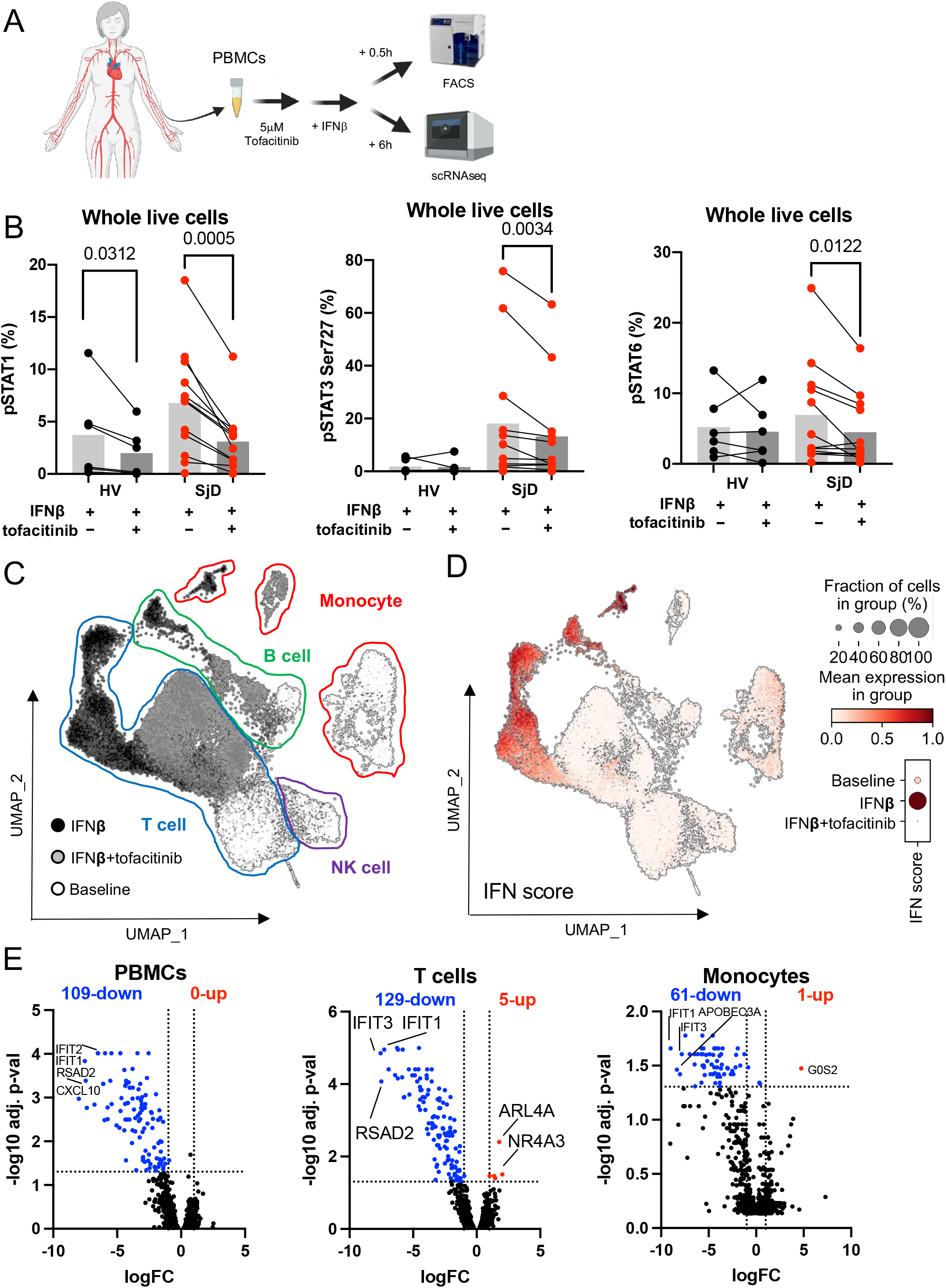
Treatment effects of tofacitinib in PBMCs. (A) Overview of using PBMC treated by tofacitinib for scRNAseq and FACS. PBMCs were treated with or without 5mM tofacitinib for 1 hour prior to IFNβ treatment for 30 minutes or 6 hours, respectively. (B) Treatment with tofacitinib blocked STATs phosphorylation status induced by IFNβ stimulation in PBMCs from SjD. P value was calculated using Mann-Whitney test. (C) UMAP embedding of the entire dataset colored by generated clusters labelled by 4 general cell type annotations. (D) Differential utilization of IFN signature of each condition showed tofacitinib abolished the IFNβ-induced IFN score to baseline level. (E) Volcano plot showing DEGs by pseudobulk analysis across SjD and HV, in which some representative genes were highlighted. T cells (129 genes) and monocytes (61 genes) exhibited greater numbers of downregulated ISGs in the context of tofacitinib. ISGs (e.g., *IFIT1*, *IFIT3*) were among the top DEG in all subsets after IFNβ and tofacitinib normalizing this response. RNAseq, RNA sequencing; SjD, Sjogren’s Disease; DEG, differentially expressed genes; HV, healthy volunteer; ISG, interferon stimulated genes.

### Tofacitinib, a JAK inhibitor, reduces Type-I IFN response in primary salivary gland epithelial cells

To model the glandular epithelial response to Type-I IFN and blockade by tofacitinib, we used primary salivary gland epithelial cells derived and cultured from SjD and HV. Tofacitinib at 5μM concentration did not induce apoptosis or necrosis in pSGEC (**supplemental figure 7A, B**). Tofacitinib mitigated IFNβ-induced pSTAT1 proteins in both the nucleus and cytosol in pSGECs as assessed by IF (0.78-fold, *p<0.0001*; 0.84-fold, *p<0.0001*, respectively; **figure 7B**), and dose-dependently decreased IFNβ-induced ISGs mRNA expression (*CXCL10*: 0.002-fold, *p=0.023*; *ISG15*: 0.03-fold, *p=0.003*; *MX1*: 0.01-fold, *p=0.009*; **figure 7C** and **supplemental figure 7C**). Of note, SjD-derived pSGECs exhibited an enhanced response to IFNβ stimulation (e.g., *CXCL10*: 33.6-fold, *p=0.032*; **figure 7C**). pSGECs treated with tofacitinib without IFNβ stimulation showed a trend towards decreased expression of ISGs, though this was not statistically significant (**supplemental figure 7D**). These results suggest that *i*) responsiveness to JAK inhibition is activation-dependent and SjD-selective, and *ii*) while activation is not retained in pSGECs *in vitro*, pSGECs do retain SjD-specific enhanced responsiveness to IFN stimulation^20^ supporting the robustness of this model system.

**Figure 7:**
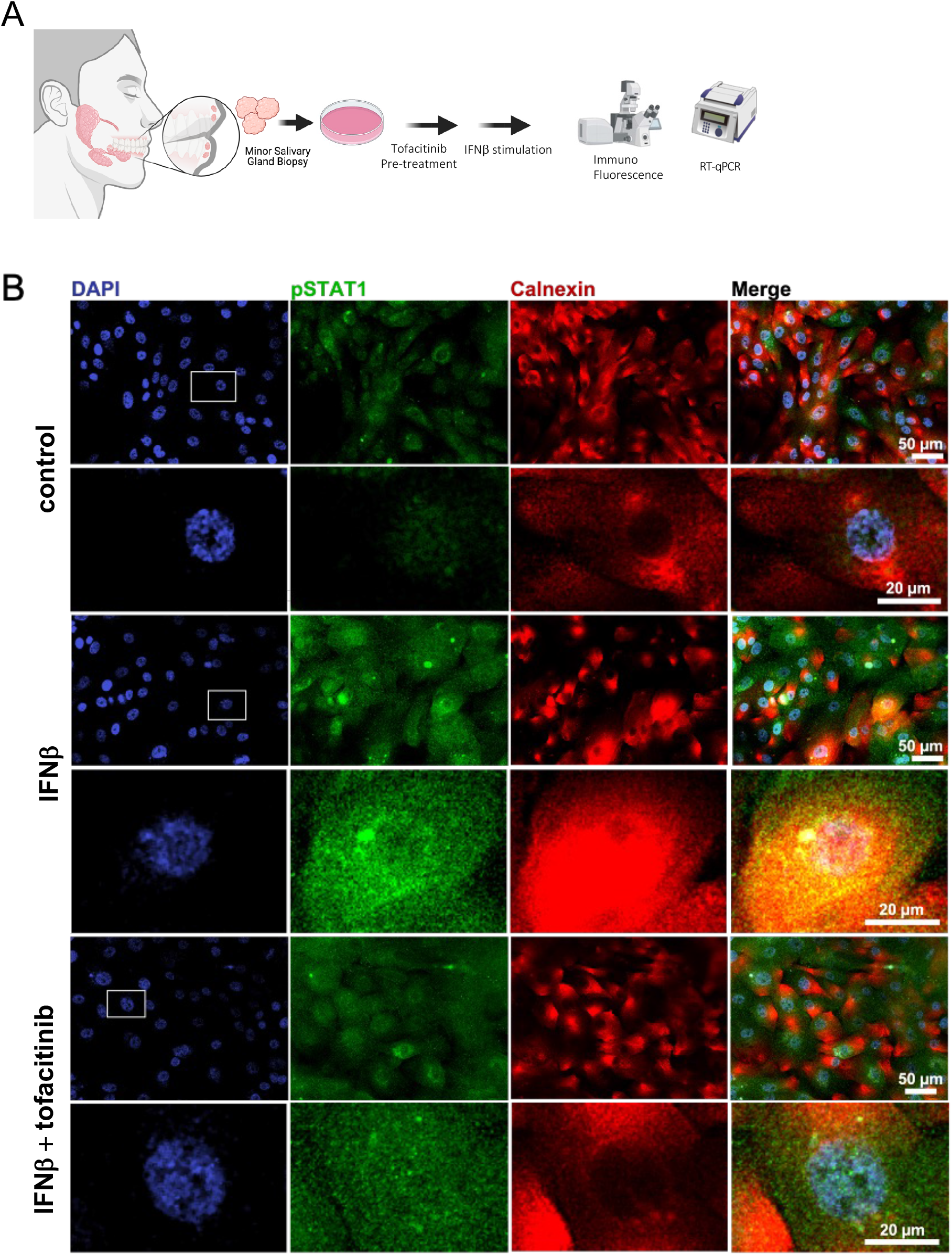
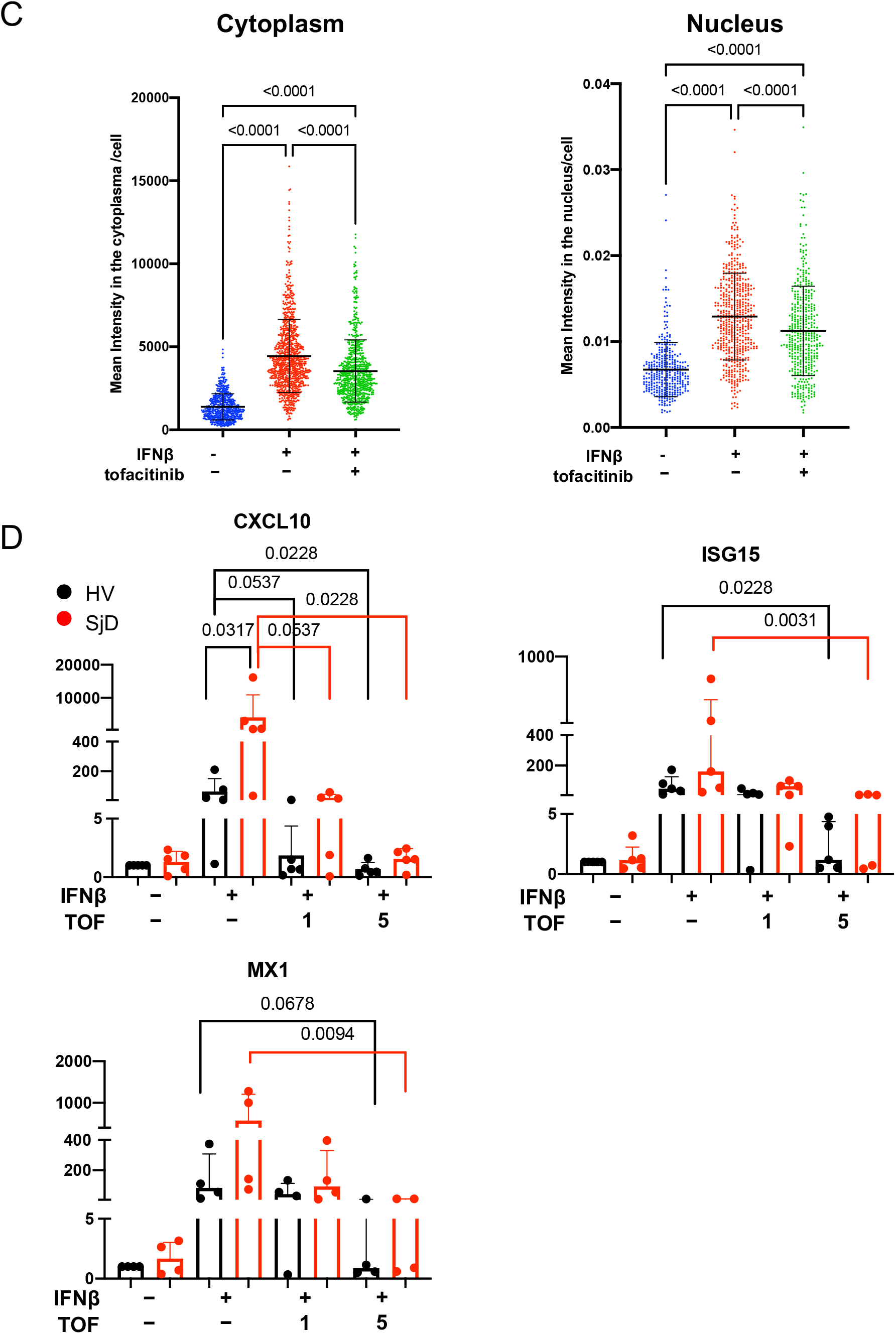
Treatment effects of tofacitinib in pSGECs. (A) Overview of using pSGEC treated by tofacitinib for IF and RT-qPCR. pSGEC were derived from fresh MSG biopsies. (B, C) Differential expression of pSTAT1 in pSGECs. Tofacitinib treatment mitigated IFNβ-induced STAT1 phosphorylation in both the nucleus and cytosol. P value was calculated using Kruskal-Wallis test. (D) Expression change of ISGs (*i.e., CXCL10*, *ISG15*, *MX1*) on pSGECs showing dose-dependently decreased IFNβ-induced ISGs mRNA expression. 5 SjD and 5 HV (n=5 individuals’ lines, respectively). 1 SjD and 1 HV samples were eliminated from MX1 result. P value was calculated using Mann-Whitney test and Kruskal-Wallis test. pSGEC, primary salivary gland epithelial cells; SjD, Sjogren’s Disease; HV, healthy volunteer; ISG, interferon stimulated genes.

## Discussion

Current strategies for managing SjD are ineffective in limiting disease progression or preventing the long-term decline in quality of life. Recent successes of early stage randomized controlled trials in SjD (BAFF-R, TACI, CD40, CD40L^21^) offer renewed optimism to patients and providers. Given the multiple intersecting pathogenic pathways driving varied clinical presentations in SjD, and a paucity of successful trials, there is continued need for biologically relevant, effective, and targeted therapeutics. Rational repurposing of approved drugs offers an exciting path to accelerate the approval of safe and effective medications for patients with SjD. To this end, we established a multimodal testing platform using human tissues from SjD patients to identify actionable targets and to directly test biological effects. Specifically, we hone-in on Type-I IFN signaling through the JAK-STAT pathway.

The activation of the IFN pathway in SjD patients’ peripheral blood has been reported^22^, but the specific cell types driving this upregulation in blood or in the glands remain poorly detailed. Using transcriptomic and functional approaches on peripheral blood and MSG tissue from SjD patients, our data revealed enhanced IFN signaling via JAK-STAT in both compartments, emphasizing the therapeutic potential of this pathway. We identified cell subsets driving this activated state: monocytes, NK cells and DCs in PBMCs and infiltrating immune cells and salivary gland epithelial cells in MSGs. Our data confirm SjD-specific upregulation of ISGs and JAK-STAT pathway in salivary glands and blood, aligning with previous reports.^23^ These cytokines, via Janus Kinases, initiate and enhance inflammation in SjD salivary glands.^2,24–28^ Similar to a previous report, the elevated IFN signatures in MSG correlated with clinical variables including focus score and enrichment in autoantibody-positive subjects.^29^

To understand the upstream effects in SjD-affected MSG, we measured JAK genes and protein expression in the glands at single cell resolution. Among the JAK genes expressed in the MSGs, *JAK1* was most abundant and ubiquitously expressed in both SjD and non-SjD. However, JAK3 mRNA and protein expression was only elevated in SjD. Using IF on MSGs, JAK1 protein localized to immune and epithelial cells with a very slight increased expression (∼6%) in SjD immune cells likely due to increased expression in APCs as found in scRNAseq. Confirming our bulk and scRNAseq data, IF showed that JAK3 protein was increased in *i*) the ductal and acinar epithelium in SjD involved by focal lymphocytic inflammation, and *ii*) in inflammatory cells within inflammatory foci. A previous report showed JAK1 staining in ductal cells, and to a lesser extent in acinar cells, in HV and SjD patients.^30^ We extend the understanding of JAK-STAT signaling in the gland by showing the novel finding of disease-specific dependence of JAK3 signaling in the ductal epithelium. These data provide guidance for rational drug selection (i.e., a JAK3/JAK1 targeting drug such as tofacitinib) whereby the epithelium and disease-specific immune infiltrates show evidence of dependence on JAK-STAT signaling through JAK3/JAK1 in SjD.

Deciphering the JAK-STAT pathway involvement in SjD is an exciting area of investigation. We and others have demonstrated significantly increased *IL21* and IL-21-inducible genes: *JAK3* and *STAT1* in SjD via RNAseq of MSGs.^31^ pSTAT1 in SjD MSG biopsies has been associated with IFN-α, IFN-γ, and IL-6 stimulation.^32,33^ Very recently, pSTAT1 was shown to confer histopathological value in MSGs with or without lymphocytic foci^34^; however, these findings are based on weak intensity immunohistochemical staining of pSTAT1 without rigorous quantification. We now directly confirmed that pSTAT1 and pSTAT3(Ser727) are elevated in the epithelial and immune cells in the MSGs by phospho-flow cytometry from SjD subjects. It has been reported that STAT protein expression is increased in immune cells of MSG.^32,33,35^ Furthermore, our transcriptomic (bulk and sc) results show that STATs gene expressions are upregulated in SjD, and our proteomic data establishes that the levels of pSTATs are higher in SjD MSG than controls. These data further confirm that activated JAK-STAT signaling is a potentially targetable pathogenetic factor for SjD.

Our data supports that activation of IFN pathway in SjD is a systemic phenomenon and not constrained to the salivary glands.^36^ Enhanced IFN response in PBMCs could be involved in both the systemic features of SjD and local inflammation after infiltration into MSGs. T cells could proliferate locally in the salivary glands, but much of their infiltration is from migration from the circulation.^36^ In PBMCs from SjD subjects, pSTAT1 frequency was significantly higher compared to controls and was more of a general phenomenon across multiple cell subsets. On the other hand, pSTAT3(Ser727), but not –(Tyr705), were more frequently phosphorylated in SjD T cells, NK cells, and monocytes (**supplemental figure 5**). pSTAT3(Ser727) has been suggested to contribute to oxidative phosphorylation in the mitochondria with an effect that is independent of STAT3(Tyr705) phosphorylation.^37^ Davies et al.^38^ showed increased responses from B cells in peripheral blood from patients with SjD to TLR-7 and –9 agonism were dependent upon phosphorylation of STAT3 at the Ser727 site. It has been reported that Tyr705 phosphorylation is essential for STAT dimerization, nuclear translocation and DNA binding, whereas C-terminal Ser727 phosphorylation is required for maximal transcriptional activity.^39^ The increased response was found to be correlated with Type-I IFN signature suggesting that pSTAT3(Ser727) plays a key role in IFN signaling in SjD patients and could be a potential molecule for targeted therapy or to monitor therapeutic effects of targeted therapies. These results support *i*) the independence of Ser727 from Tyr705 on pSTAT3, and *ii*) possibly highlight patients with maximal pathway activation who may benefit from JAKi.

JAKi have emerged as an important new class of oral therapy for several autoimmune diseases.^40,41^ Tofacitinib was the first oral JAK inhibitor approved for rheumatoid arthritis. It blocks JAK3 and JAK1 but also has a role in JAK2 and TYK2 inhibition.^42–46^ Our *ex vivo* experiments using pSGECs and PBMCs consistently demonstrated that tofacitinib treatment suppressed JAK-STAT pathway activation in a dose-dependent manner, as shown by downregulated pSTAT levels, decreased IFN signature and ISG expression. Of note, basal elevations in pSTAT levels were selectively controlled by tofacitinib in SjD patients’ PBMCs, but not controls. Similar to our results, recently baricitinib, a semi-selective inhibitor of JAK1 and JAK2, mitigated IFN-γ-induced CXCL10 production in salivary gland ductal cell line.^30^

Cell type differences in responsiveness have been reported with regards to the potency of JAKi to inhibit cytokine signaling. A 3-fold difference was observed in the JAK1/TYK2-dependent IFN-stimulated pSTAT1 between CD4+ T cells and monocytes, whereas the potencies were comparable for B and NK cells.^47^ For the reactivity of each pSTAT, JAK inhibition of JAK1/TYK2-mediated IFNα-driven pSTAT5 and pSTAT3 was more potent than pSTAT1, potentially demonstrating the reliance on TYK2 for regulating STAT1 phosphorylation.^48^ In our study, pSTATs (e.g., pSTAT1, pSTAT3(Ser727), pSTAT6) in T cells showed consistent sensitivity to JAKi treatment. Other cell subsets also demonstrated pSTAT specific sensitivity (B cells – pSTAT1; NK cells–pSTAT1/6). Our findings suggest that pathogenic inflammatory signaling through specific JAKs may identify selective therapeutic drug choice.

Limitations of this study include relatively modest sample sizes used for fresh tissue analyses. The prospective use of fresh tissues poses logistical challenges including recruitment and adequate tissue recovery for both clinical and research purposes. Despite these limitations, we demonstrate biologically meaningful changes in JAK-STAT pathway in glandular and peripheral blood samples and direct responsiveness to JAKi. These limitations are addressed by using multiple different experimental approaches to systematically and comprehensively evaluate the IFN-JAK-STAT pathway in SjD.

In conclusion, SjD patients have increased IFN signature with activated JAK-STAT pathway which plays a key role in both the glandular and extraglandular pathogenesis of SjD. This activation is cell specific, with salivary epithelial cells and infiltrating immune cells driving the bulk of this signature in MSGs. The activated IFN-JAK-STAT signature is also seen in peripheral blood, highlighting the dysregulated systemic immune system in SjD. Modulation of the JAK-STAT pathway, through JAKi was non-cytotoxic and effective *ex vivo* using human tissues. These results suggest tofacitinib as a potential therapeutic strategy for SjD patients and serve as the basis of an open and enrolling Phase Ib/IIa randomized controlled trial to treat SjD with tofacitinib (NCT04496960).

## Data Availability

All large data sets produced in the present study will be available through dbGAP. Additional experimental data are available after reasonable request to the authors after the final publication of the manuscript, except where otherwise prohibited by patient consent groups.

https://www.ncbi.nlm.nih.gov/projects/gap/cgi-bin/study.cgi?study_id=phs001842.v1.p1

https://www.ncbi.nlm.nih.gov/projects/gap/cgi-bin/study.cgi?study_id=phs002446.v1.p1,

## Acknowledgements

The authors provide their emphatic appreciation to the subjects for their unwavering support of our clinical studies through their volunteerism. We also thank the support and staff of the NIDCR Office of the Clinical Director, NIDCR Dental Clinic, the National Eye Institute Ophthalmology Clinic, and the NCI Center for Cancer Research Anatomic Pathology.

