## Supplemental Information, Tables, Figures for "Inhibition of JAK-STAT pathway corrects salivary gland inflammation and interferon driven immune activation in Sjögren’s Disease"

**Study investigators**

**Supplemental methods**

**Histopathological Assessments**

H&E stained sections were cut (4-5mm) from formalin fixed, paraffin embedded (FFPE) MSG biopsies. All MSG sections were interpreted by the same board-certified surgical pathologist (D.E.K.) blinded to clinical classification. Salivary gland inflammation and fibrosis were graded according to Greenspan et al.[1] and Tarpley et al.[2] For MSG sections with Greenspan grade 3 or 4 sialadenitis, a focus score was calculated according to Daniels et al.[3] Additional 4-5mm sections were cut from a select set of cases for immunofluorescence microscopy.

**Human Salivary Gland RNA Sequencing**

Total RNA was isolated from minor salivary glands snap frozen in OCT using the miRCURY™ RNA Isolation Kit-Cell & Plant (Exiqon). RNA (200–500ng) was rRNA depleted with the RiboMinus Eukaryotic Kit v2 (Ambion) and subject to library preparation using the Ion Total RNA-Seq kit v2 (Thermo Fisher Scientific) according to manufacturer protocols for total RNA libraries as indicated. The barcoded cDNA library was quantified a Bioanalyzer 2100 (Agilent) and input for template preparation using the Ion PI™ Hi-Q™ Chef Kit and the Ion Chef instrument, followed by sequencing on the Ion Proton sequencer (Thermo Fisher Scientific). Bulk RNA sequencing was performed as previously described . RNAseq data that passed quality control was deposited in dbGaP: phs001842.v1.p1.

**Single Cell RNA Sequencing of the MSG and PBMCs**

***Tissue dissociation.*** Seven SjD and five non-SjD subjects provided MSG biopsies and samples were processed for scRNAseq as previously described (online supplemental table 1).[4 5] After excision, MSGs (2-3/per patient) were placed immediately in ice-cold RPMI. MSGs were placed in a sterile 100 mm tissue culture dish and delicately dissected into uniform ~1 mm lobules. Lobules were dissociated using the Miltenyi Multi-tissue Dissociation Kit A using the Multi_A01 in C-type tubes at 37 °C in an OctoMACS tissue disruptor using heated sleeves. Single-cell suspensions were filtered through 70- and 30-μm filters and rinsed with 1× Hanks' buffered salt supplemented with 1% ultrapure, DNase/RNase-free, bovine serum albumin solution. Cells were centrifuged at 300*g* for 10 min at 4 °C and washed twice with 1× Hanks' buffered salt solution. Cell counting and viability were determined using a Trypan blue exclusion assay. Suspensions with greater than 75% viability were used for subsequent sequencing. In all instances, adequate numbers of glands were submitted for histopathological assessment and standardized focus scoring as described above.

***Single-cell capture, library preparation and sequencing.*** Single-cell suspensions targeting approximately 10,000 cells were prepared as described above and loaded onto a 10x Genomics Chromium Next GEM Chip B (10x Genomics) following the manufacturer's recommendations.

After cell capture, single-cell library preparation was performed following the instructions for the 10x Chromium Next GEM Single Cell 3' kit v3 (10x Genomics). The libraries were pooled and sequenced on four lanes of a NextSeq500 sequencer (Illumina), adopting the read configuration indicated by the manufacturer.

***scRNAseq data processing, quality control, and analysis.***

Read processing was performed using the 10x Genomics workflow (10x Genomics). Briefly, the Cell Ranger v3.0.1 Single-Cell Software Suite was used for demultiplexing, barcode assignment and UMI quantification (http://software.10xgenomics.com/single-cell/overview/welcome). Sequencing reads were aligned to the hg38 reference genome (Genome Reference Consortium Human Build 38) using a pre-built annotation package obtained from the 10x Genomics website (https:// www.10xgenomics.com/). Samples were demultiplexed using the 'cell ranger mkfastq' function, and gene count matrices were generated using the 'cellranger coun' function.

***Single-cell RNA sequencing data was analyzed in Python using Scanpy.***

Cells containing less than 100 genes and genes expressing in less than 10 cells were filtered out. Raw data was normalized as count per ten thousand and then logarithmized. Individual sample libraries were combined and processed as a single library using the Batch balanced kNN procedure.[6] Cell clustering was performed by the Leiden graph-clustering method[7] and displayed in Uniform Manifold Approximation and Projection (UMAP) format. Type I IFN score was calculated based on the average expression of 21 Type I IFN stimulated genes (ISGs) using the "scanpy.tl.score_genes" command.[8] Differentially expressed genes (DEGs) were identified using the Wilcoxon model in the "scanpy.tl.rank_genes_groups" function after excluding ribosomal and mitochondrial genes. Enriched pathway analysis in DEGs was performed using DAVID Bioinformatics Resources (https://david.ncifcrf.gov).

Cells containing more than 200 and fewer than 2,500 unique features were retained. From this set, cells with greater than 15% of read counts attributed to mitochondrial DNA were filtered out. We adjusted this value from 5% to 15% to increase the yield from each sample and did not observe substantive changes in our results after adjustment.

**Immunofluorescence microscopy and analysis on MSG**

***Detection of JAK1 and JAK3 in FFPE biopsies of minor salivary glands.***

Sections (5 μm) from FFPE MSG were baked for 1hrs at 65°C, deparaffinized and rehydrated following standard procedures. Antigen retrieval was performed using R-UNIVERSAL Epitope Recovery Buffer (1x) (Electron Microscopy Sciences) in microwave pressure cooker using 80% power for 15 min. Sections were blocked for 30 min RT using 10% FBS buffer in PBS plus saponin 0.01%, and incubated in diluted primary antibodies overnight at 4 °C. Primary antibodies were washed in PBS for 10 min, 3 times, and a mix of secondary antibodies were applied in blocking buffer (online supplemental table 2). After 45 minutes of incubation, secondaries antibodies were rinsed, and tissues were mounted using mounting solution. Whole slides were digitally scanned using the 40X objective using an Axioscan Z1 (Zeiss), with the following exposure times (DAPI, 20ms; JAK1, 200ms; JAK3, 150ms; and Cytokeratin-18, 50ms).

Whole slide images were uploaded into Visiopharm Image Analysis software V2022.11 (Visiopharm A/S). Digitized tissue sections were identified, cells were detected and segmented using deep-learning-based nuclear segmentation, cellular phenotyping was assessed using the PhenoApp® module using thresholds based on visual assessment of positivity. PhenoApp® was trained on all the analyzed samples. After establishment of cellular phenotypes, the median fluorescence intensity per cell was normalized based on the background autofluorescence per slide and the max and min intensity in the set. Dimensionality reduction using *t*-Stochastic Nearest-neighbor Embedding (*t*SNE) was used to cluster segmented cells and visualize individual cellular phenotypes based on protein expression of JAK1, JAK3, and Cytokeratin-18.

***Immunofluorescence in primary cell culture assays.***

Primary salivary gland epithelial cells (pSGECs) were generated according to methods published by Jang et al.,[9] from patients fulfilling ACR 2016 classification criteria or HV.[10] pSGECs were plated on chamber slides and stimulated as indicated. The cells were fixed for 2h at 4°C and permeabilized for 10 min at −20°C. Cells were stained with primary antibodies overnight and then stained by secondary antibody for 2h (online supplemental table 2). Images were acquired on Nicon A1 HD (Nicon) confocal microscope and processed with CellProfiler in ImageJ (Broad Institute).[11]

***Assessment of serum proteome***

Proteomic profiles were measured in serum (50uL) using the SOMAscan Assay V1.3 (SomaLogic, Inc.) at the Trans-NIH Center for Human Immunology, Autoimmunity, and Inflammation (CHI), National Institutes of Health (Bethesda, MD, USA) as previously reported.[12] Following the manufacturer's data processing guidelines, the raw data were normalized by control hybridization, then median signal normalization and finally, inter-plate calibration normalization. Once normalized, the data levels of protein were compared. The type I IFN protein (IFNP) scores were calculated based on Smith et al.[13] Prior to calculation of the IFNP, the four proteins characterized in the IFNP were $log_2$ transformed and scaled to the mean and standard deviation of the respective sample distribution. Data processing was performed in -R 3.1.1.

**Flow cytometry**

*Minor Salivary Gland Flow Cytometry.* Freshly biopsied minor salivary glands were dissociated and enumerated as described above. Cells were fixed using BD Cytofix Fixation Buffer (BD) containing 4.2% formaldehyde, washed with staining buffer, then permeabilized with BD phosflow Perm Buffer Ⅲ (BD). Multicolor flow cytometry was used to quantify the phosphorylation status of each pSTAT in gated cell subset populations (online supplemental table 2 and online supplemental method 1). Cells were acquired using a FACS Symphony (BD Biosciences) flow cytometer and analyzed with FlowJo ™ v10.8 (BD Life Sciences).

*Peripheral blood mononuclear cells.* Flow cytometry analysis was performed using cryopreserved peripheral blood mononuclear cells (PBMCs) isolated by BD Vacutainer® CPT™ Cell Preparation Tube (BD Biosciences) for basal or experimental analyses. Thawed PBMCs were stimulated as indicated in 500 µl media in 48-well flat-bottom plates at 37°C, 5% $CO_2$ and 95% humidity according to experimental conditions. Cells were treated as described above and analyzed (online supplemental table 2 and online supplemental method 2).

Multicolor flow cytometry was used to quantify the phosphorylation status of pSTATs in gated cell subset populations (supplemental table 2 and supplemental method 1,2). Cells were acquired using a FACS Symphony (BD Biosciences) flow cytometer and analyzed with FlowJo™ v10.8 (BD Life Sciences).

**RNA isolation and RT-qPCR**

RNA was isolated using Monarch Total RNA miniprep Kit (New England BioLabs). Standard Taqman assays (supplemental table 3) were performed in technical triplicates with at least biological duplicates to measure relative gene expression using the delta-delta Ct method[14] on a QuantStudio™ 6 Pro Real-Time PCR System (Thermo Fisher).

**Western Immunoblot Analysis**

Extracted total protein was resolved using sodium dodecyl sulfate-polyacrylamide gel electrophoresis (SDS-PAGE) (4% stacking, 12% resolving, Invitrogen) and transferred onto Polyvinylidene fluoride (PVDF) membranes. Then, membranes were incubated with primary and secondary antibodies (online supplemental Table 2). The signal was detected by ChemiDoc^MP Imaging System (BIO-RAD), and the density of the bands was analyzed using Fiji.[15]

***LDH assay and AnnexinV staining***

The potential for cytotoxicity or apoptosis induced by tofacitinib was measured using the plate-based CyQUANT™ LDH Cytotoxicity Assay kit (Invitrogen) and the flow-cytometry-based Dead Cell Apoptosis Kit with Annexin V Alexa Fluor 488 & Propidium Iodide (Invitrogen) following the manufacturer's protocol on a FACS Symphony flow cytometer.

#### *Statistical Analysis for Difference in Immune Cell Populations/Cytokines*

Statistical methods were employed using GraphPad Prism (GraphPad), matlab, or -R as described, and the type and nature of the data were considered when assessing differences in mean values and variances across biological and experimental replicates. Generally, a *p*-value of <0.05 was considered statistically significant, unless otherwise noted (e.g., where adjusted *p*-values to compensate for multiple comparisons). Specific statistical methods are reported in specific methodological sections and/or figure legends. For the analysis of pseudobulk RNAseq from scRNAseq data, the *voom* pipeline from the *limma* package (Bioconductor) was used determine DEG from the pseudo-bulk scRNAseq expression profile of individual clusters with cut-offs of adjusted *p*-value of <0.05 and fold-change >2-forld or <0.5-fold expressed.

### Supplemental Method 1: Gating strategy of MSG

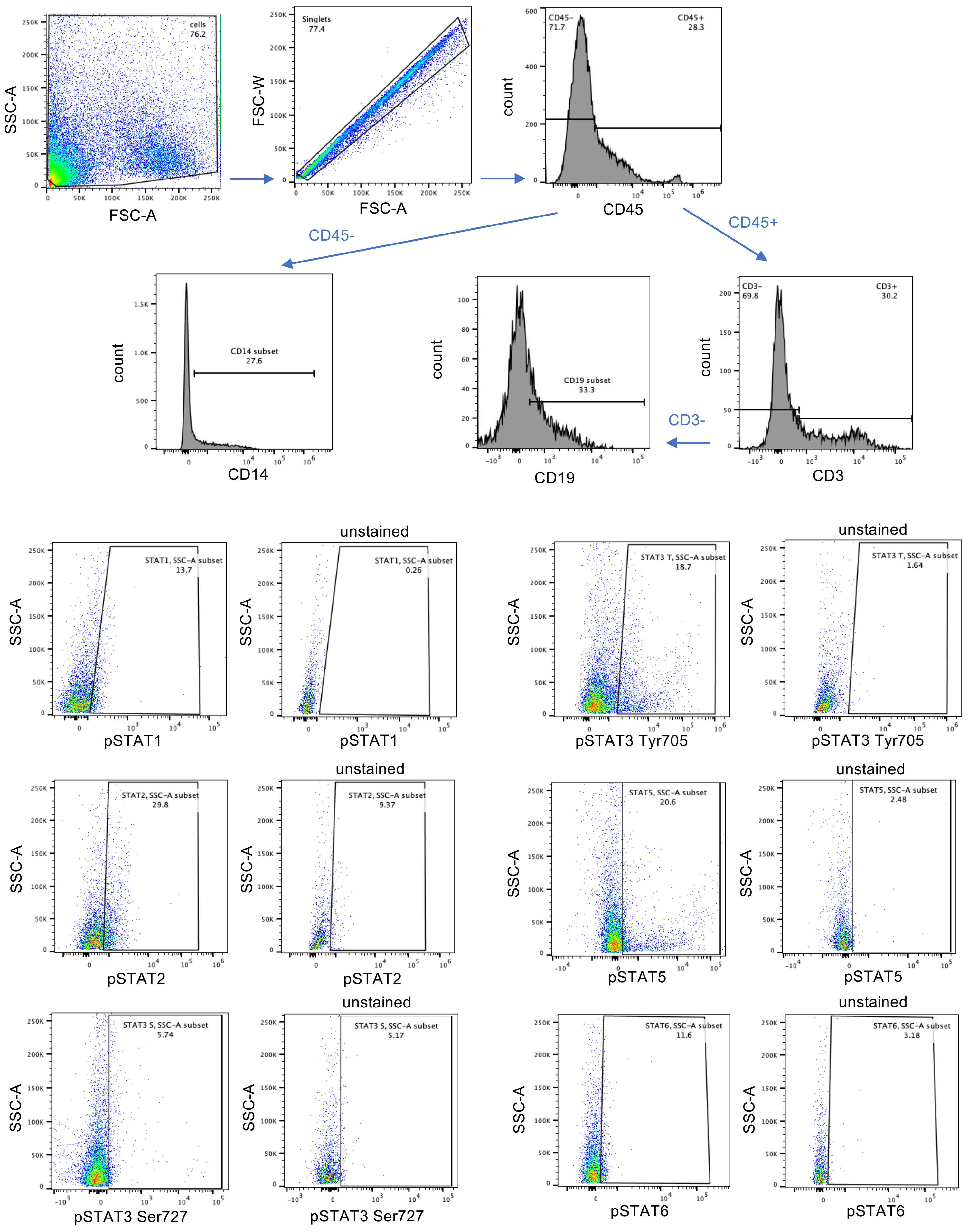

### Supplemental Method 2: Gating strategy of PBMC

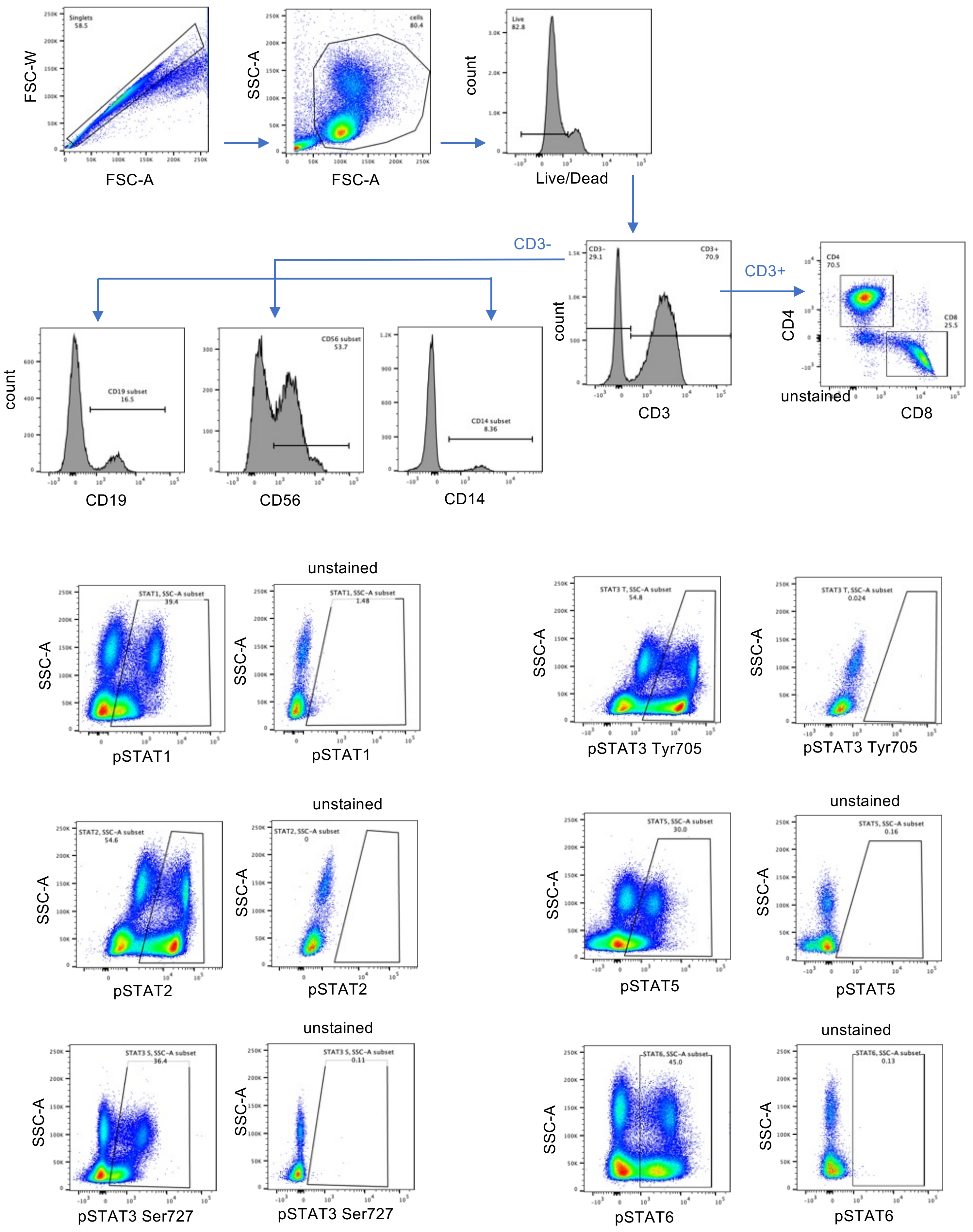

**Supplemental Figure 1: Bulk sequencing of minor salivary gland and IFN signature**
(A) Differential expression of IFN score in SjD and HV. Kruskal-Walls test. (B) Gene graph enrichment analysis showed consistent and direct regulation of the JAK-STAT pathway signaling through *IL7/IL15/IL21* via *JAK3* (solid red connections). (C) Differential expression of JAK-STAT related genes in SjD and HV. (D) Differential expression of *IFNG* showing increased *IFNG* in SjD (~3-fold, *p*=0.0258) and the correlation between Type I and II sum of Z-scores. P value was calculated using Mann-Whitney test and Spearman correlation.

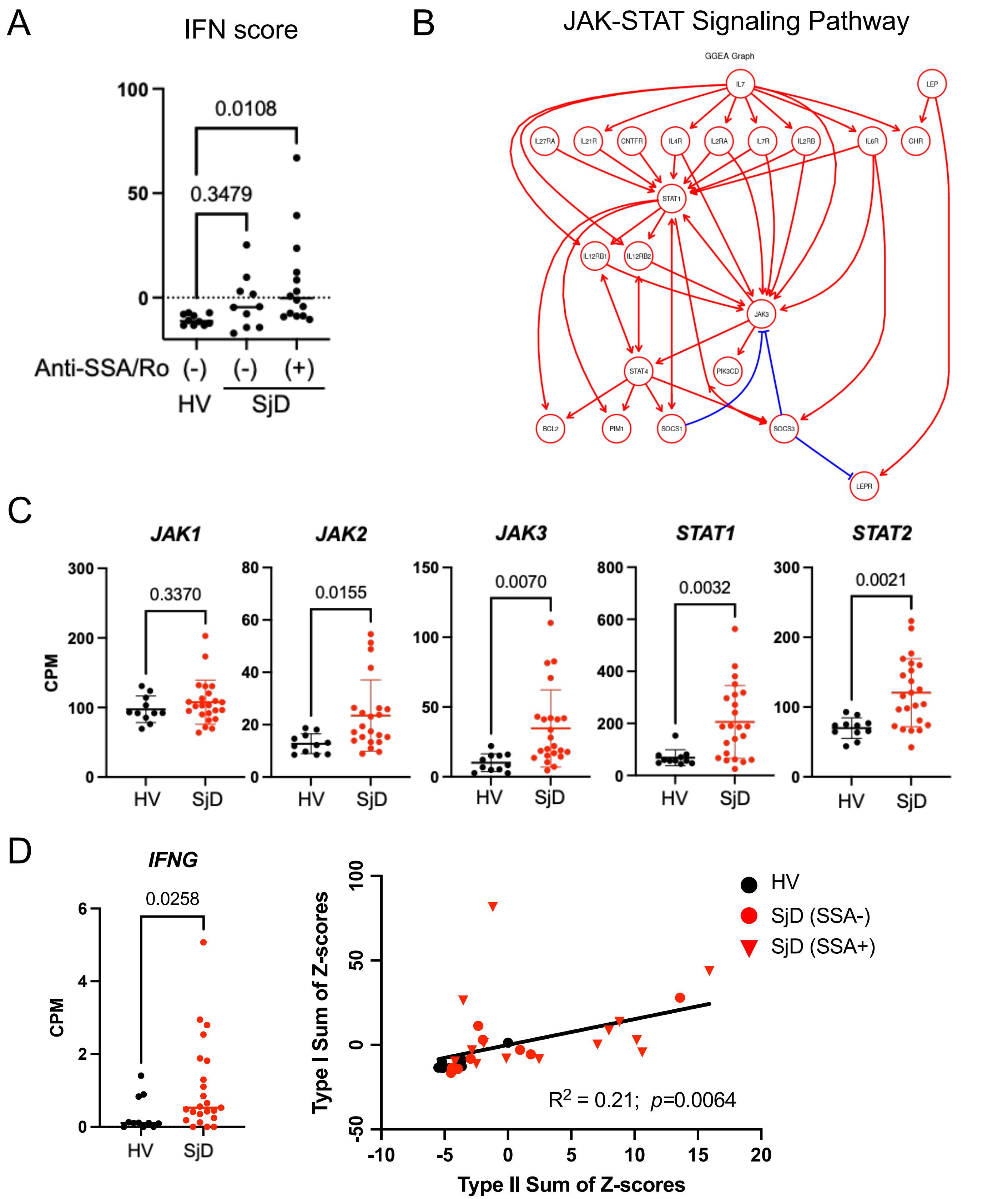

**Supplemental Figure 2: immunofluorescence microscopy**

(A, B) Unbiased segmentation and quantitation of per cell expression in SjD and non-SjD. (C) Cell subset population difference in SjD and non-SjD MSG from IF. The cellular proportion was changed to  less epithelial and more immune cells in the SjD glands. (D) tSNE figures in SjD and non-SjD MSG from IF. JAK1 and JAK3 expressions were clearly elevated in immune cells in SjD, but both expression change in the SjD epithelial cells were mild.

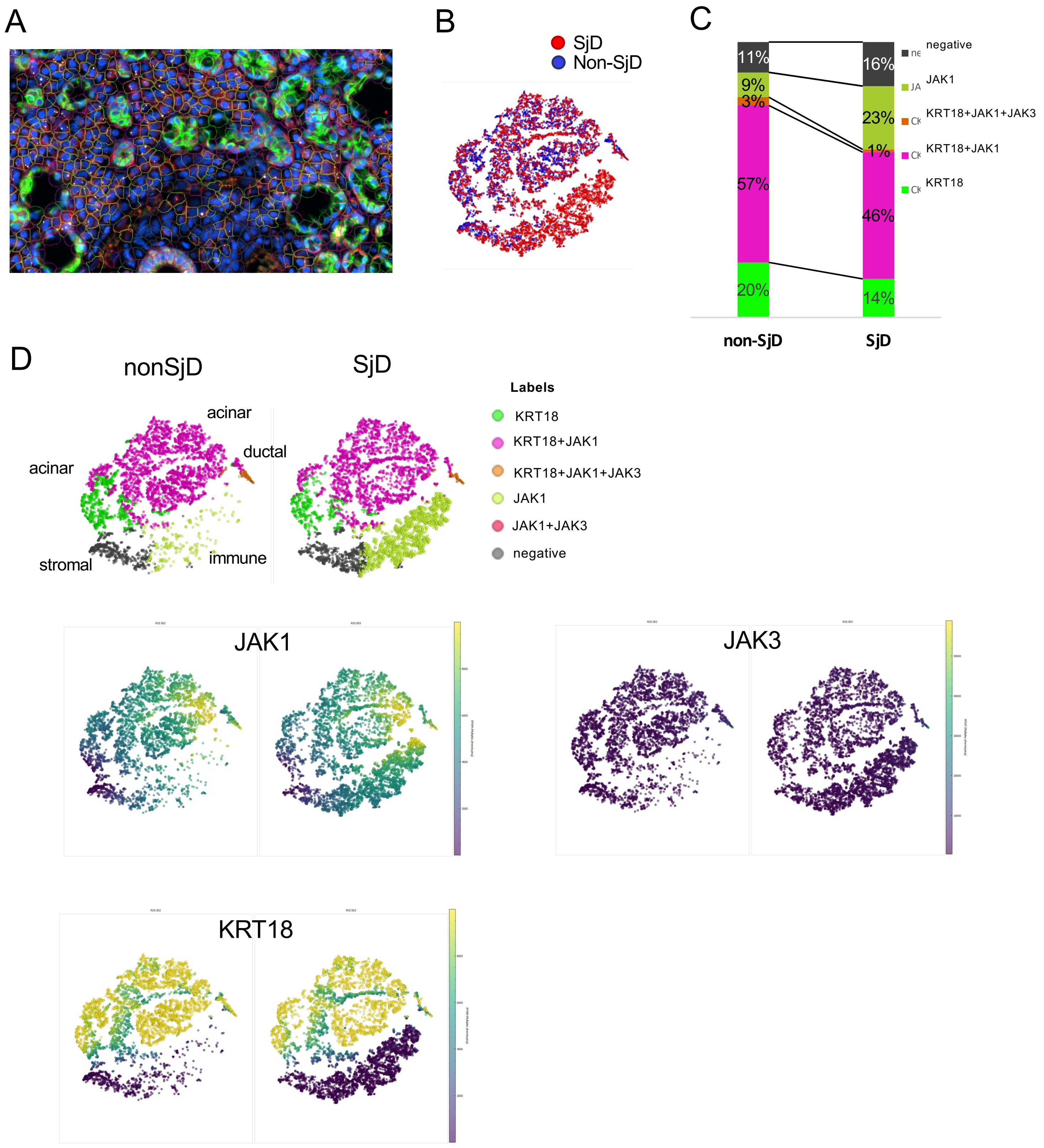

**Supplemental Figure 3: Single cell RNAseq of MSG**

(A-D) Differential gene expressions in SjD and non-SjD MSG. *JAK1* was the most ubiquitously expressed in MSGs with appreciably less expression of *JAK2*, *JAK3*, and *TYK2*. Seromucous acinar cells showed increased expression of all *JAKs*, while ductal cells had increased expression of *JAK3* and *TYK2;* APCs and plasma cells exhibited increased expression of *JAK1*, *JAK3*, *TYK2. STATs* gene expression showed increased expression of *STAT1* in all cell clusters in MSGs from patients with SjD, while others were expressed in only specific cell clusters.

A

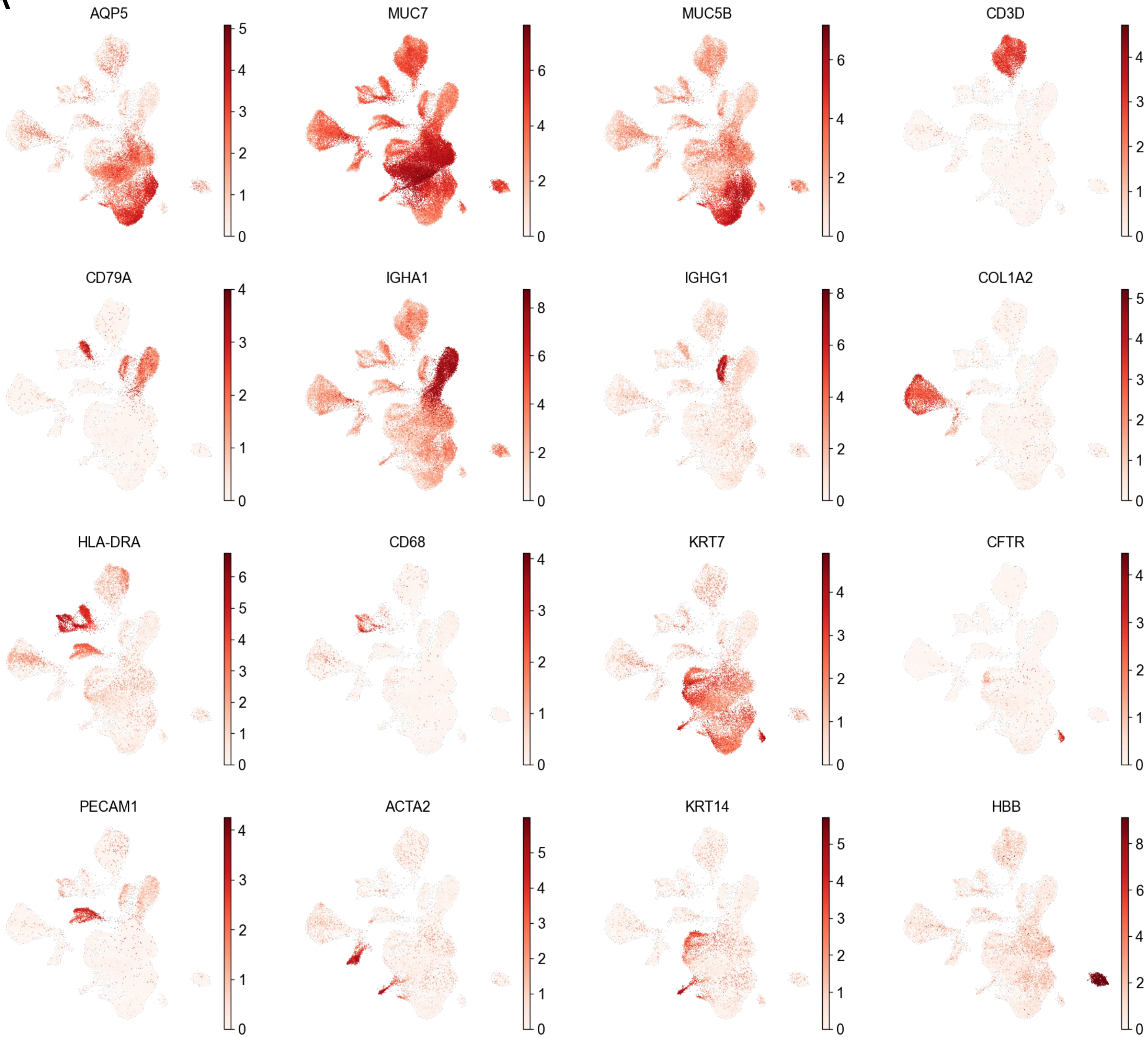

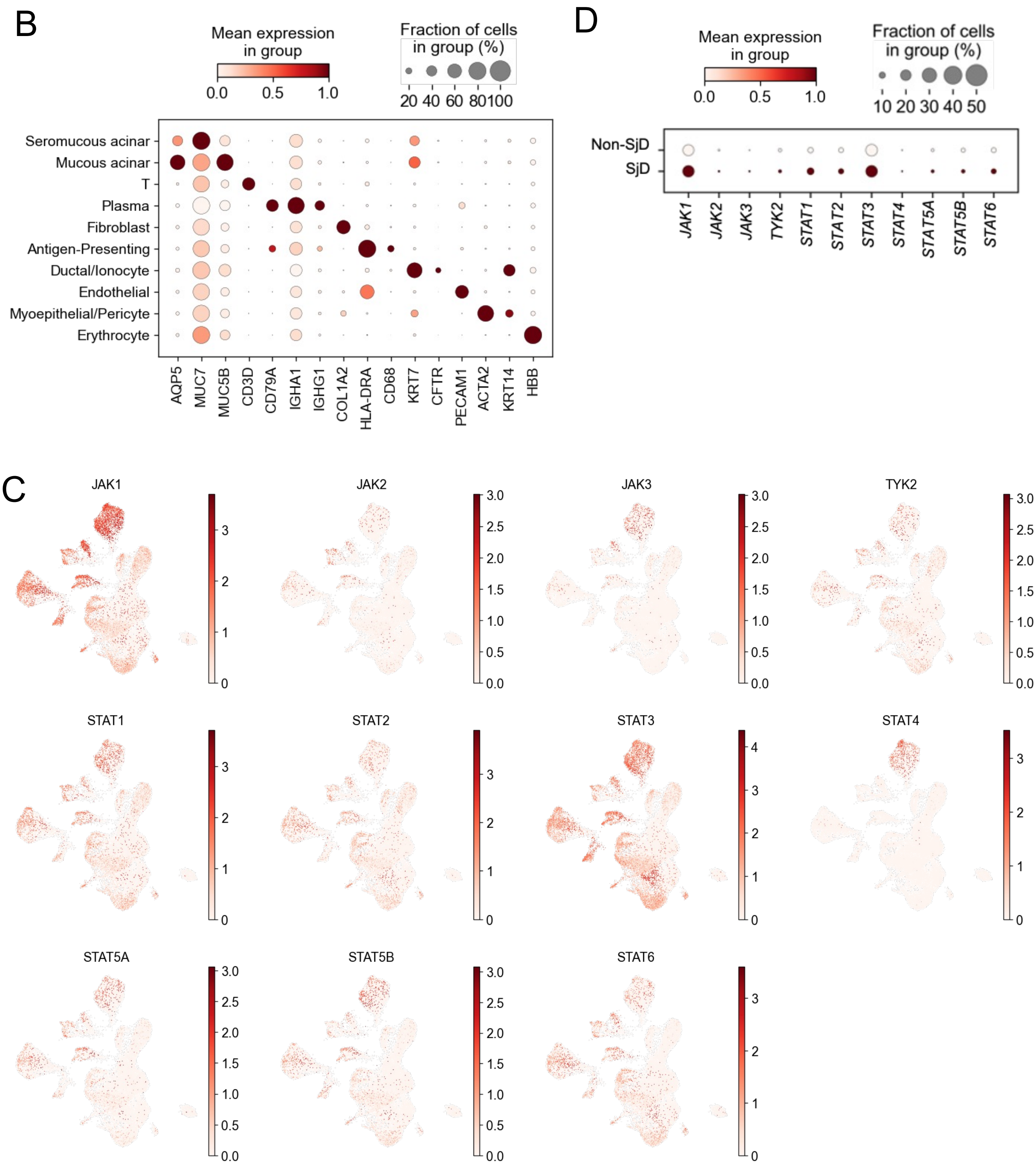
B
Mean expression in group
0.0 0.5 1.0
Fraction of cells in group (%)
20 40 60 80100
Seromucous acinar
Mucous acinar
T
Plasma
Fibroblast
Antigen-Presenting
Ductal/Ionocyte
Endothelial
Myoepithelial/Pericyte
Erythrocyte
AQP5 MUC7 MUC5B CD3D CD79A IGHA1 IGHG1 COL1A2 HLA-DRA CD68 KRT7 CFTR PECAM1 ACTA2 KRT14 HBB
D
Mean expression in group
0.0 0.5 1.0
Fraction of cells in group (%)
10 20 30 40 50
Non-SjD
SjD
JAK1 JAK2 JAK3 TYK2 STAT1 STAT2 STAT3 STAT4 STAT5A STAT5B STAT6
C
JAK1
JAK2
JAK3
TYK2
STAT1
STAT2
STAT3
STAT4
STAT5A
STAT5B
STAT6

**Supplemental Figure 4: Single cell RNAseq of PBMC**
(A-C) Differential gene expressions in SjD and non-SjD MSG. DEG analysis showed upregulation of many ISGs (e.g., *IFI44L, IFIT3, ISG15, MX1,* and *IFI6*) in SjD PBMCs.

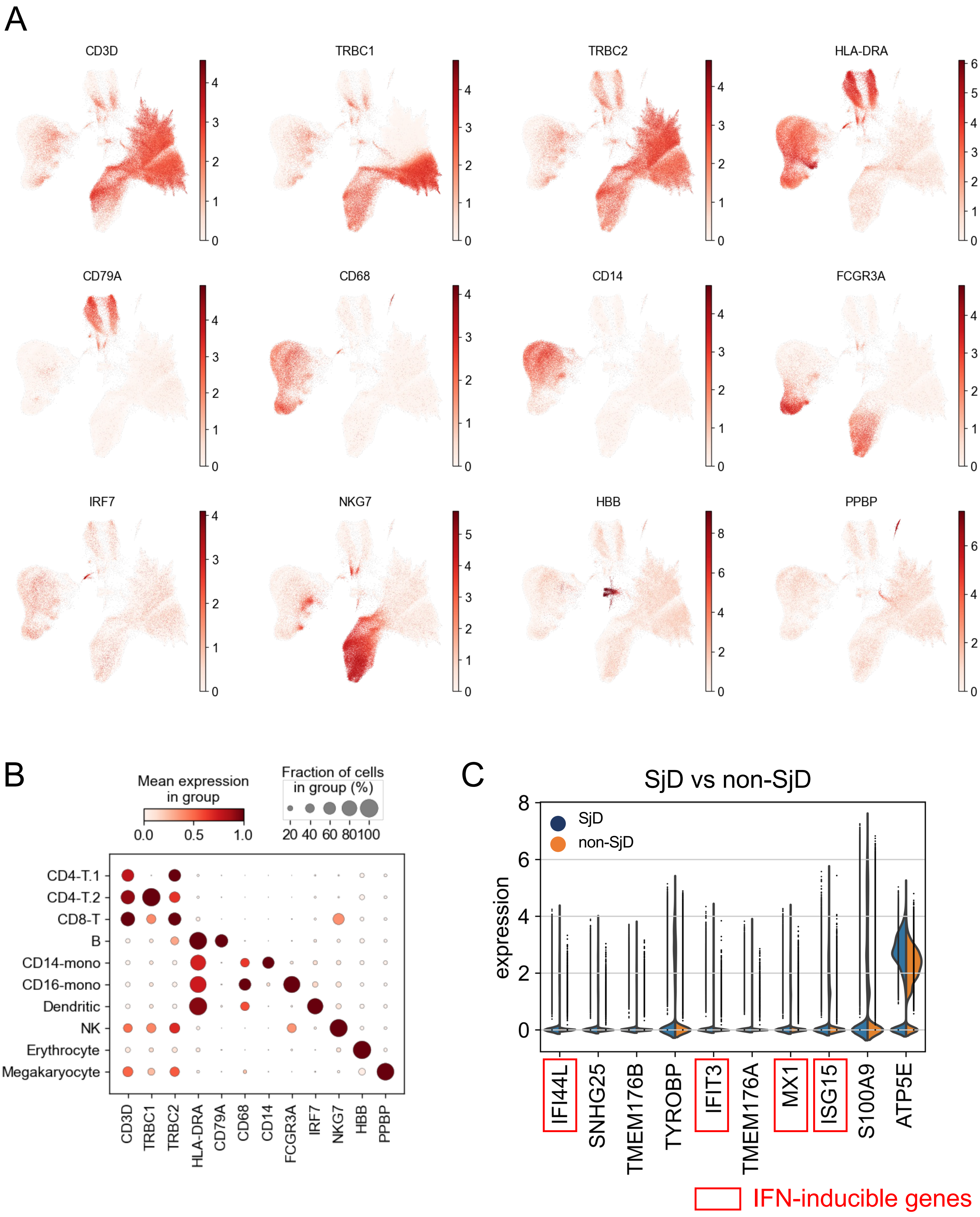

**Supplemental Figure 5: Basal pSTATs frequencies in PBMCs**
(A-D) Basal pSTAT frequencies in SjD and HV PBMCs. The frequency of pSTAT1, pSTAT3(Ser727), and pSTAT6 were higher in SjD patients compared to HV, but not for pSTAT3(Tyr705).
* p<0.05, **p<0.01, P value was calculated using Welch's test.

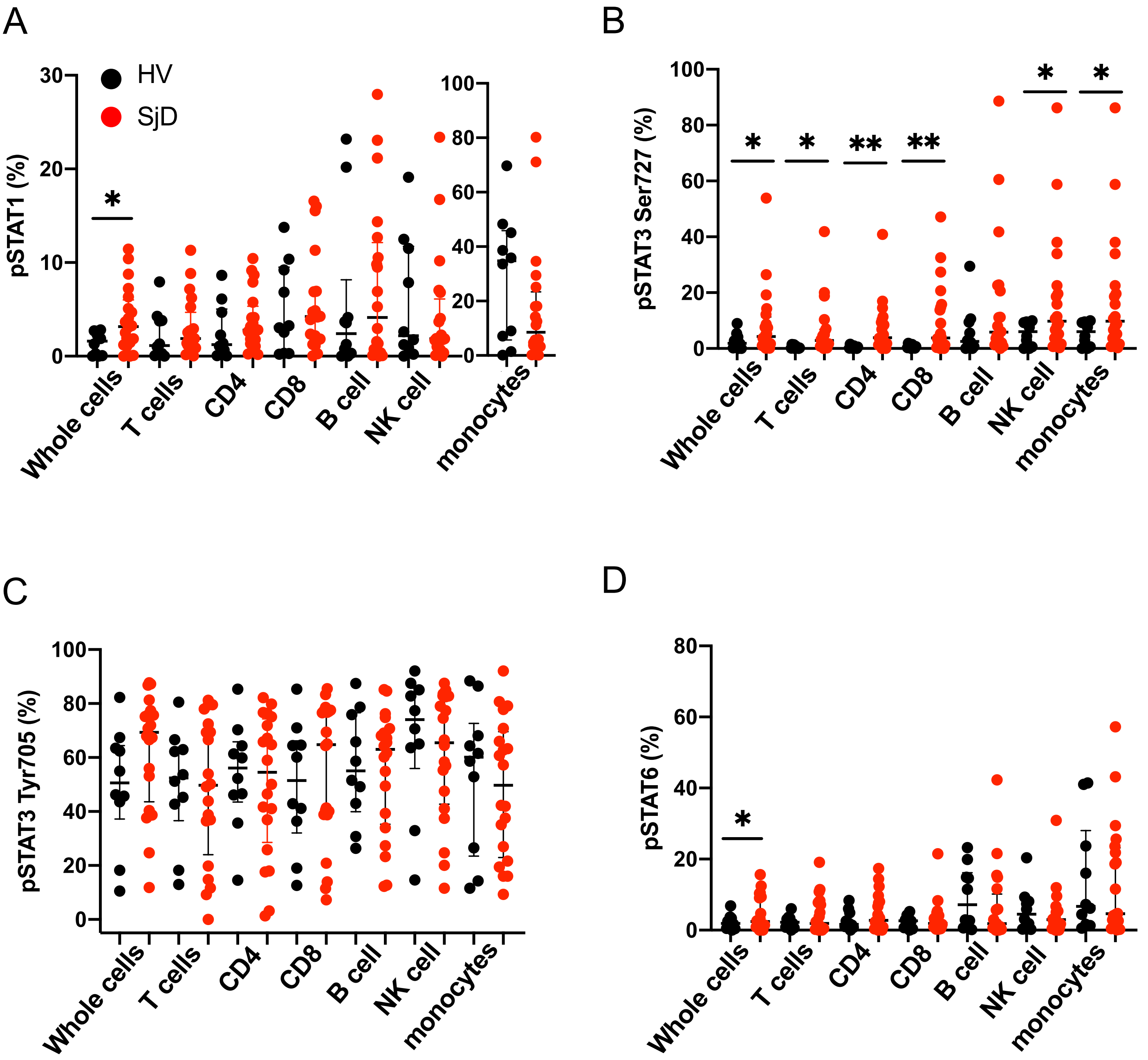

**Supplemental Figure 6: Treatment effects of tofacitinib in PBMCs**
(A, C) Effects of tofacitinib on with or without IFN-β induced pSTATs in PBMCs. Tofacitinib treatment significantly downregulated pSTAT levels in SjD. This trend was not seen in HV which showed lower levels even without tofacitinib treatment. P value was calculated using Mann-Whitney test. (B) Tofacitinib abolished the IFNβ-induced IFN score to baseline level on other cell subsets. (D) 5mM of tofacitinib did not induce necrosis in PBMC under these experimental conditions.

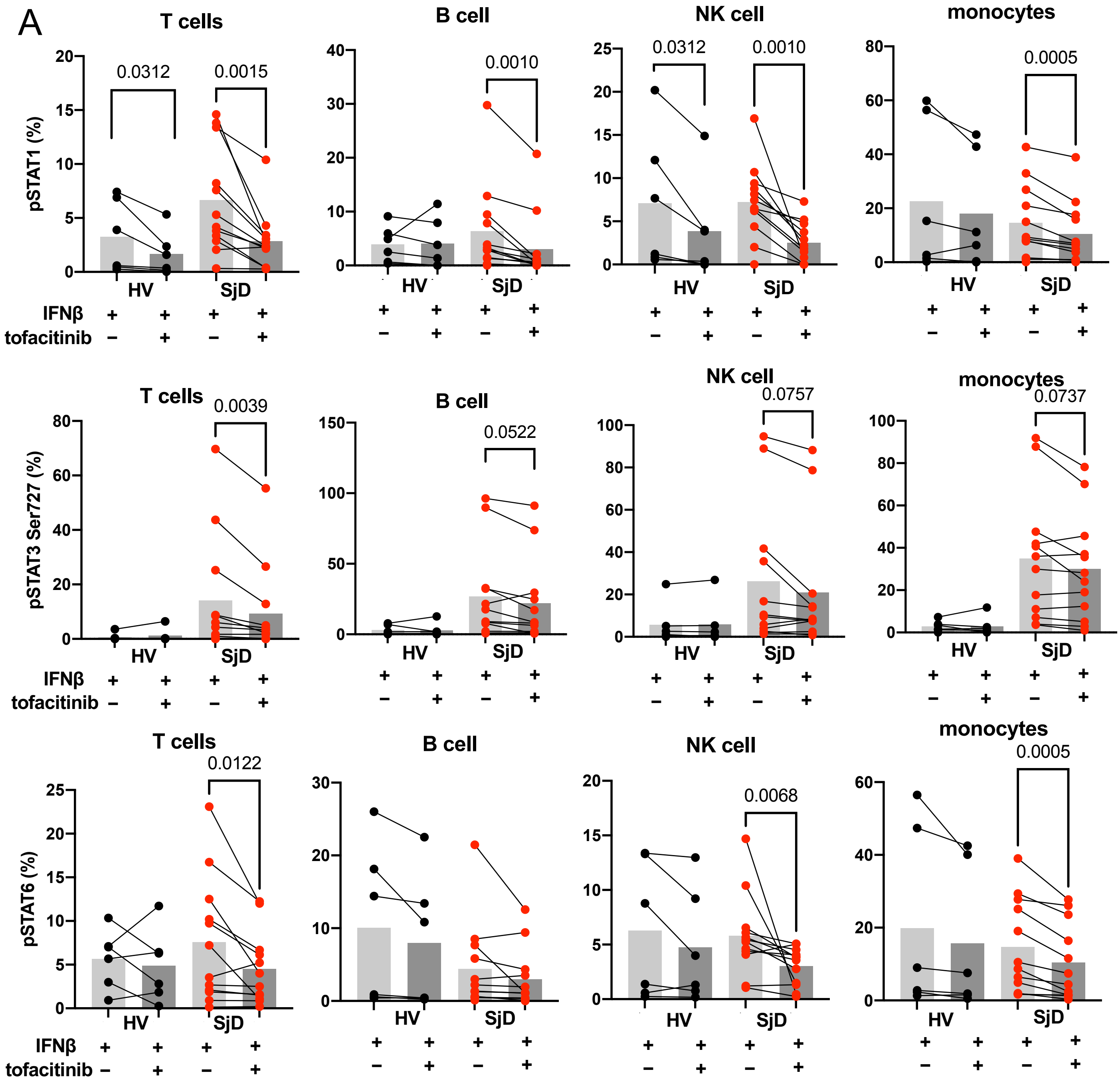

B

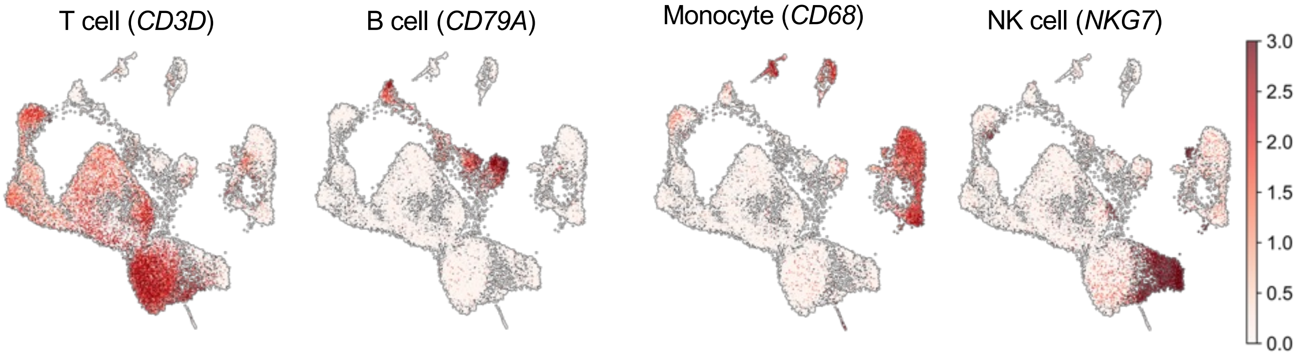
T cell (CD3D)
B cell (CD79A)
Monocyte (CD68)
NK cell (NKG7)
3.0
2.5
2.0
1.5
1.0
0.5
0.0

C

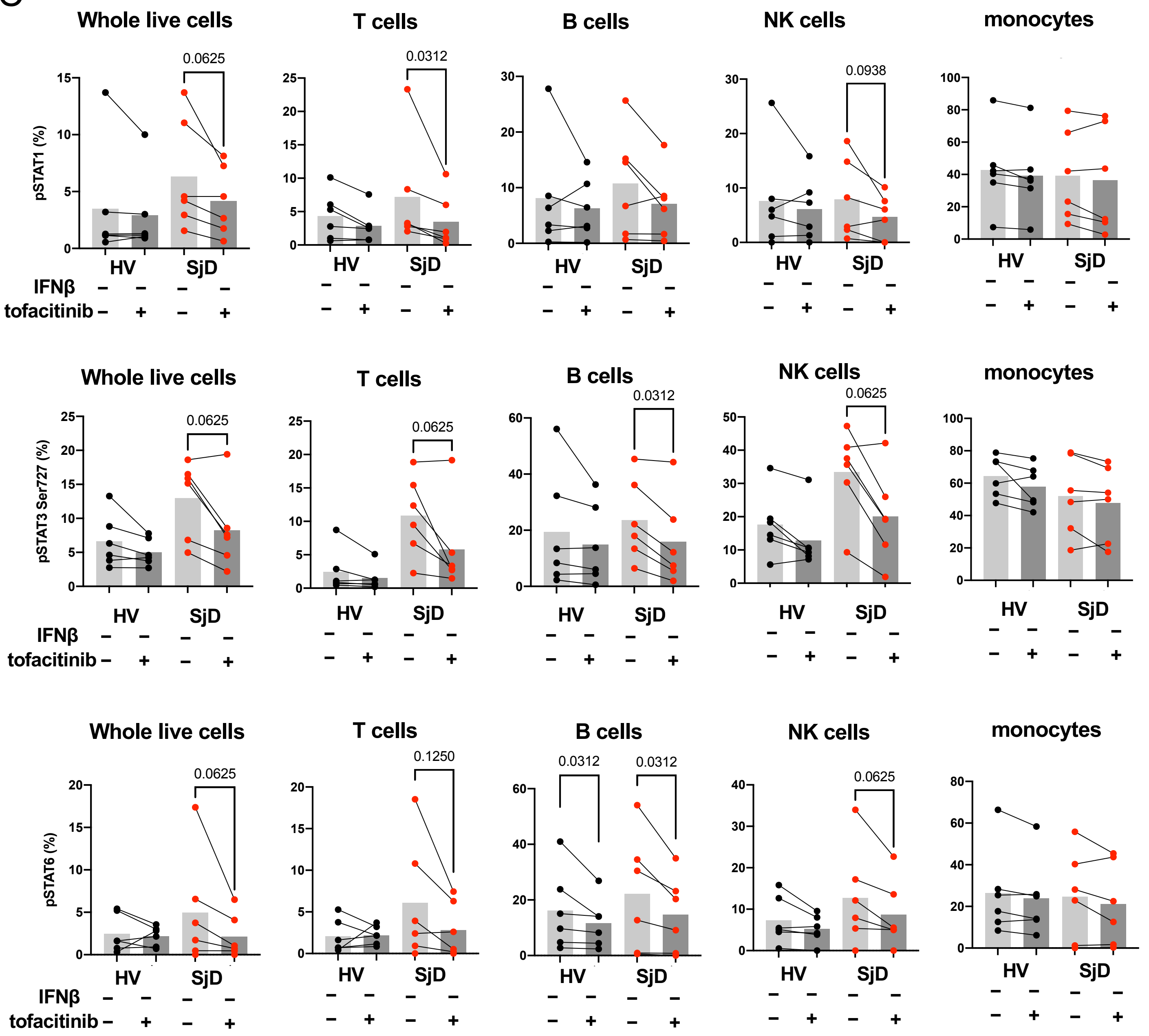
Whole live cells
T cells
B cells
NK cells
monocytes
pSTAT1 (%)
0.0625
0.0312
0.0938
pSTAT3 Ser727 (%)
0.0625
0.0625
0.0312
0.0625
pSTAT6 (%)
0.0625
0.1250
0.0312
0.0312
0.0625
HV
SjD
IFNβ
tofacitinib

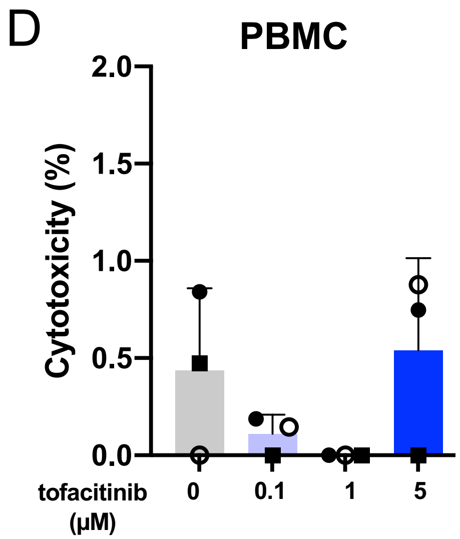
D
PBMC
Cytotoxicity (%)
2.0
1.5
1.0
0.5
0.0
tofacitinib (μM)
0
0.1
1
5

**Supplemental Figure 7: Treatment effects of tofacitinib in pSGECs**
(A, B) 5µM of tofacitinib did not induce apoptosis and necrosis in pSGEC under these experimental conditions.
(C, D) Effects of tofacitinib on with or without IFN-β induced pSTATs in pSGEC.

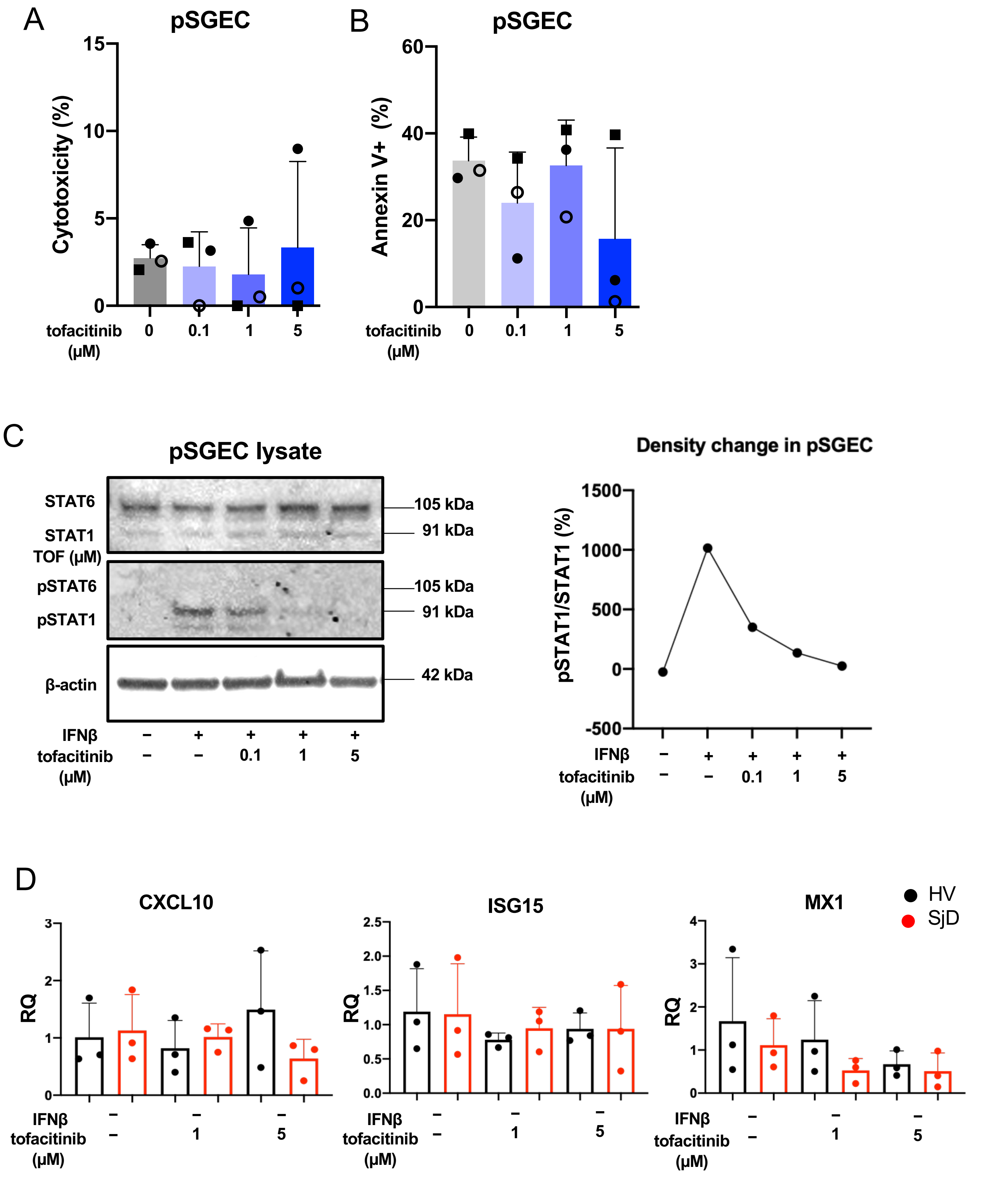

**Supplemental Table 1: Cohort description**

Descriptions of the cohorts used for each analysis. For race, values are numbers of white/black/Asian or other study subjects included in each analysis. Significant differences in age in the cohort are in comparison to the HV or SjD cohort. * *p*<0.05, P value was calculated using Mann-Whitney test. UK: unknown.

| Analysis | Subject group | Sample size (gender) | Age median (range) | Race | Salivary flow (mL, range) | FS (range) |
|---|---|---|---|---|---|---|
| Bulk RNAseq of MSGs (Figure 1) | HV | 11 (M1 F6 UK4) | 30 (24-57) | 3/2/5 | 13.9 (7.4-22.5) | – |
| | SjD | 23 (M0 F21 UK2) | 52 (27-75) | 8/6/9 | 3.8 (0-16.4) | 3 (1-12) |
| Basal pSTAT levels in MSG (Figure 2) | non-SjD | 6 (M1 F5) | 41.5 (34-75) | 4/1/2 | 16.2 (0.4-72.3) | – |
| | SjD | 7 (M1 F6) | 50 (41-69) | 3/3/1 | 6.7 (0-27.8) | 1 (1-6) |
| scRNAseq of MSGs (Figure 3) | non-SjD | 5 (M2 F3) | 48 (39-56) | 5/0/0 | 8.3 (0.8-19.5) | – |
| | SjD | 7 (M1 F6) | 55 (43-75) | 3/4/0 | 1.9 (0-13.2) | 2 |
| Somalogic in serum (Figure 4) | HV | 32 | – | – | – | – |
| | non-SjD | 50 | – | – | – | – |
| | SjD | 78 | – | – | – | – |
| scRNAseq of PBMCs (Figure 4) | non-SjD | 6 (M2 F4) | 51.5 (39-61) | 5/1/0 | 8.4 (2.9-16.3) | – |
| | SjD | 8 (M1 F7) | 50.5 (43-69) | 5/2/1 | 7.8 (0-14.3) | 1.5 (1-3) |
| Basal pSTAT levels in PBMC (Figure 5) | HV | 10 (M0 F10) | 42 (20-85) | 8/1/1 | 18.1 (6.2-44.0) | – |
| | SjD | 20 (M0 F20) | 57 (36-78)* | 11/5/4 | 9.5 (0-55.1) | 3 (0-12) |
| Tofacitinib treatment in PBMC (Figure 6) | HV | 6 (M0 F6) | 42 (25-85) | 6/0/0 | 17.3 (6.2-43) | – |
| | SjD | 12 (M0 F12) | 56 (45-69) | 5/3/4 | 9.5 (0-28.1) | 4 (0-12) |
| RT-qPCR (Figure 7) | HV | 5 (M0 F6)) | 43 (23-62) | 4/1/0 | 20.4 (2.24-44.0) | – |
| | SjD | 5 (M1 F5) | 63 (61-81)* | 2/2/1 | 1.0 (0-23.1) | 3 (3-5) |

### Supplemental Table 2: Antibodies and dilution

| Antibody | | Manufacturer | Dilution |
|---|---|---|---|
| **Flow Cytometry** | | | |
| Cell surface staining | LIVE/DEAD™ Fixable Aqua Dead Cell Stain Kit | Invitrogen | 1:100 |
| | APC-Cy7-labelled anti-mouse CD3 antibody | BioLegend | 1:100 |
| | PerCP/Cy5.5-labelled anti-mouse CD4 antibody | BioLegend | 1:100 |
| | BV510-labelled anti-mouse CD8 antibody | BioLegend | 1:100 |
| | BV605-labeled anti-mouse CD19 antibody | BioLegend | 1:100 |
| | BV711-labeled anti-mouse CD56 | BioLegend | 1:100 |
| | BV421-labeled anti-mouse CD14 | BioLegend | 1:100 |
| Intracellular staining | Phospho-Stat1 (Tyr701) (58D6) Rabbit mAb (PE Conjugate) | Cell Signaling Technology | 1:100 |
| | Phospho-Stat2 (Tyr690) (D3P2P) Rabbit mAb (Alexa Fluor® 488 Conjugate) | Cell Signaling Technology | 1:100 |
| | Phospho-Stat3 (Tyr705) (D3A7) XP® Rabbit mAb (Alexa Fluor® 488 Conjugate) | Cell Signaling Technology | 1:100 |
| | Phospho-Stat3 (Ser727) (D4X3C) Rabbit mAb (Alexa Fluor® 647 Conjugate) | Cell Signaling Technology | 1:100 |
| | Phospho-Stat5 (Tyr694) (D47E7) XP® Rabbit mAb (Alexa Fluor® 555 Conjugate) | Cell Signaling Technology | 1:100 |
| | Phospho-Stat6 (Tyr641) (D8S9Y) Rabbit mAb (Alexa Fluor® 647 Conjugate) | Cell Signaling Technology | 1:100 |
| | Cytokeratin 18 Antibody (LDK18) Mouse mAb (DyLight405 Conjugate) | NOVUS Biologicals | 1:100 |
| **Western Blots** | | | |
| HRP-conjugated anti β-actin Mouse mAb | | SIGMA | 1:10000 |
| STAT1 (9H2) Mouse antibody | | Cell Signaling Technology | 1:500 |
| Phospho-Stat1 (Tyr701) (D4A7) Rabbit antibody | | Cell Signaling Technology | 1:500 |
| STAT6 Mouse antibody (177C322.1) | | Novus Biologicals | 1:500 |
| Phospho-Stat6 (Tyr641) Rabbit antibody | | Cell Signaling Technology | 1:500 |
| IRDye® 800CW Donkey anti Rabbit IgG (H + L) | | LI-COR Biosciences | 1:10000 |
| IRDye® 680RD Donkey anti-Mouse IgG (H + L) | | LI-COR Biosciences | 1:10000 |
| SuperSignal™ West Pico PLUS Chemiluminescent Subtrate | | Thermo Scientific | |
| **Immunofluorescence confocal microscopy** | | | |
| ProLong™ Diamond Antifade Mountant with DAPI | | Invitrogen | - |
| Fig.2 | mouse anti-JAK1 (GTX34019) | Gene Tex | 1:100 |
| | rabbit anti-JAK3 (GTX34020) | Gene Tex | 1:100 |
| | goat anti-Cytokeratin 18 (LS-B11232-50) | LsBIO | 1:200 |
| | Alexa Fluor® 550 Donkey anti-mouse | Jason Immuno Research | 1:300 |
| | Alexa Fluor® 647 Donkey anti-rabbit | Jason Immuno Research | 1:300 |
| | Alexa Fluor® 488 Donkey anti-goat | Jason Immuno Research | 1:300 |
| Fig.6 | Anti-CANX goat mAb | SICGEN | 1:100 |
| | Phospho-Stat1 (Y701) rabbit mAb | abcam | 1:100 |
| | Alexa Fluor® 594 AffiniPure Donkey Anti-Goat IgG (H+L) | Jason Immuno Research | 1:300 |
| | Fluorescein (FITC) AffiniPure F(ab')$_2$ Fragment Donkey Anti-Rabbit IgG (H+L) | Jason Immuno Research | 1:300 |
| | Phalloidin Conjugated CF®640R | Biotium | 1:50 |

### Supplemental Table 3: Reagents for RT-qPCR

| Reagent | Manufacturer |
|---|---|
| **Standard Taqman assays** | |
| TaqMan™ Gene Expression Cells-to-CT™ Kit | invitrogen |
| TaqMan™ Fast Advanced Master Mix | invitrogen |
| TaqMan™ Gene Expression Assay (FAM) ID:Hs01060665_g1 Gene Symbol:ACTB Dye Label and Assay Concentration:FAM-MGB / 20X | invitrogen |
| TaqMan™ Gene Expression Assay (FAM) Unit size:M (750 reactions/750 µL), made to order Assay ID:Hs00171042_m1 Gene Symbol:CXCL10 Dye Label and Assay Concentration:FAM-MGB / 20X | invitrogen |
| TaqMan™ Gene Expression Assay (FAM) Unit size:S (750 reactions/750 µL), inventoried Assay ID:Hs01921425_s1 Gene Symbol:ISG15 Dye Label and Assay Concentration:FAM-MGB / 20X | invitrogen |
| TaqMan™ Gene Expression Assay (FAM) ID:Hs00895608_m1 Gene Symbol:MX1 Dye Label and Assay Concentration:FAM-MGB / 20X | invitrogen |
